# Can a General-Purpose Coding Agent Analyze a Production Hospital Data Warehouse?

**DOI:** 10.64898/2026.09.02.26362008

**Authors:** Nicholas P. Marshall, William Haberkorn, Joshua Faulkenberry, Ganga Palakkattil, Fateme Nateghi Haredasht, Hayden T. Schwenk, Jonathan H. Chen, Keith Morse

**Author notes:** **Corresponding author:** Nicholas P. Marshall, MD, 453 Quarry Road, Mail Code 5660, Palo Alto, California, 94304, USA.

## Abstract

**Background:** Health systems answer most questions by having expert analysts hand-write queries against a complex electronic health record data warehouse, a slow, resource-intensive process. Whether an autonomous coding agent can do this accurately is unknown.

**Methods:** In a single-center quality-improvement evaluation, we posed ten questions about a common pediatric infection, acute otitis media. The questions went to analysts, whose adjudicated answers were the reference, and to an autonomous coding agent (OpenAI Codex), which wrote and ran read-only queries on a full copy of the production data warehouse (Epic Caboodle). We ran the agent under four conditions: an autonomous baseline (each question answered three times at two reasoning-effort settings), a variant in which it listed its assumptions, interactive analyst feedback, and reuse of a corrected definition across related questions. Outcomes were accuracy, patient-level agreement (F1), reproducibility, and efficiency (time and tokens).

**Results:** Working autonomously, the agent wrote valid queries and never fabricated data, but rarely produced the exact answer. At medium effort, it came within 5% of the reference on 27 of 30 runs but exact on only 10. Higher effort produced no improvement. Reproducibility was the greater weakness, with the three runs returning an identical answer on only 3 of 10 questions. When a run matched the reference, it had found the same patients (F1 = 1.00); the exception was an over-counted procedure (F1 = 0.72). With analyst feedback on four questions, it answered two exactly, and reusing the corrected definition on related questions restored reproducibility and accuracy.

**Conclusions:** A general-purpose coding agent matched an adjudicated analyst reference on most routine questions but not reproducibly, because key definitions depended on warehouse knowledge that the data dictionary omits. Supplying that knowledge, by prompt or feedback, restored reproducibility. Under expert analyst supervision, the agent is already a capable drafting aid and a promising step toward broader hospital analytic support.

## 1 Introduction

Almost every operational, quality, and research question a health system asks of its own data is answered the same way: an expert analyst hand-writes a query, in Structured Query Language (SQL), against an expansive electronic health record (EHR) data warehouse. These analyses inform decisions at every level, from local patient care to nationally reported quality measures. Analyst capacity is limited, however, and demand routinely exceeds it, making these analyses a rate-limiting step for research, quality improvement, and operations. Autonomous coding agents now inspect an unfamiliar environment, write and run their own programs, and continue without step-by-step direction. Whether that capability extends to healthcare analytics is unknown.

Large language models are now common in the EHR but almost entirely in low-autonomy roles such as documentation and message drafting; 31.5% of US hospitals reported using EHR-integrated generative AI in 2024, largely for these tasks.[1] Whether an autonomous agent can write and run the queries that answer institutional questions is unanswered, and caution is warranted. On realistic schemas, general text-to-SQL still trails humans: GPT-4 reached 54.89% execution accuracy on the BIRD benchmark against 92.96% for humans,[2] and the best agent solved only 21.3% of enterprise tasks on Spider 2.0.[3] Model-drafted clinical phenotyping queries have been too broad or too narrow and required expert correction.[4]

The field has advanced largely by constraining the agent. Published clinical systems use fixed tool catalogs, sandboxes, read-only access, and mandatory human review,[5] and a survey of more than 200 healthcare agent studies identified no fully autonomous deployments.[6] The closest precedents each stop short of the analyst’s task. They include autonomous SQL agents on public research databases,[7, 8] a hospital agent reaching a real warehouse only through fixed retrieval tools,[9] function-calling agents on EHR-derived data in a simulated server,[10] an enterprise assistant limited to retrieval and predefined automations,[11] a pipeline built on curated study definitions,[12] and an agent making clinical decisions in a simulated record rather than querying a warehouse.[13] We found no peer-reviewed report of a general-purpose coding agent that writes and runs its own queries, open-endedly, against a production data warehouse and is benchmarked against expert analysts.

In this pilot, we gave a general-purpose autonomous coding agent, not a purpose-built clinical tool, read-only access to a full copy of our institution’s production EHR data warehouse and asked it to answer a graded set of analytic questions of the kind our analysts handle in routine practice, each by writing and running its own queries. Our objective was to measure how accurately and reproducibly it answered against an adjudicated analyst reference standard, whether analyst interaction improved its answers, and what oversight was required to make it usable.

## 2 Methods

This was a single-center quality-improvement and feasibility evaluation at Stanford Medicine Children’s Health, using existing operational data from ambulatory pediatric primary care clinics. The activity was determined to be quality improvement through institutional review, with Chief Quality Officer approval, and was determined by the institutional review board not to constitute human subjects research (Protocol #88354). Before any protected health information was accessed, the agent was reviewed and approved by our institutional Artificial Intelligence Oversight Committee under a business associate agreement and privacy review. Agent output did not influence clinical care. We report following SQUIRE 2.0,[14] TRIPOD-LLM,[15] and STARD 2015 items for the comparison against a reference standard.[16]

Our analysts do not query the transactional EHR (Epic Chronicles), where patient care occurs and heavy queries degrade performance. They work instead from a dedicated reporting warehouse (Epic Caboodle) that consolidates operational EHR data with external sources into a reporting-optimized model of approximately 46,000 tables. For the study, the agent queried a full copy of this warehouse in a separate instance, refreshed nightly so study queries could not affect operations. All tasks were bounded to a fixed historical window (index encounters in 2023 to 2024, follow-up through 2025), so the nightly refresh did not change the data across a task’s replicate runs; runs and scoring were performed in July and August 2026 against a reference standard finalized on July 31, 2026.

We evaluated ten analytic tasks of escalating complexity, from a single-table count to treatment failure (Table 1). Acute otitis media (AOM) was the exemplar, chosen for its frequency and capacity to generate graded questions; the study concerns analytic capability, not the disease. NPM and HTS derived the ten questions and code sets from an ongoing institutional quality-improvement project, so they reflect analyses our analysts are routinely asked to run. Each task also had a definition, the exact inclusion criteria, code sets, and date windows specifying what to count. Each task was a self-contained prompt in a fresh session, so no task could influence another, and identical prompts went to the agent and the analysts (Supplement B).

**Table 1.** Study Task Set.

| # | Task | Question | Output | Complexity |
| --- | --- | --- | --- | --- |
| 1 | Birth Month | Unique patients born in a given month (whole database) | Count | Low |
| 2 | AOM Cohort | Patients under 26 with an ambulatory AOM encounter diagnosis | Cohort | Low to Moderate |
| 3 | Recurrent AOM | AOM patients with 3 or more episodes within 6 months | Cohort | Moderate |
| 4 | Prescribed Antibiotic | AOM patients given a systemic antibiotic within 3 days of diagnosis | Cohort | Moderate |
| 5 | Antibiotic Frequency | How often each of five common antibiotics was prescribed | Counts | Moderate |
| 6 | Antibiotic Spectrum Classification | Proportion of treated patients whose first AOM antibiotic was narrow-, broad-, or other-spectrum | Proportion | Moderate |
| 7 | Tubes (Recurrent) | Recurrent-AOM patients who received tympanostomy tubes | Cohort | High |
| 8 | Median Days to Tube | Median days from first AOM to first tube | Scalar | High |
| 9 | Complications | AOM patients with any specified complication, overall and by type | Cohort | High |
| 10 | Treatment Failure | Antibiotic change on days +2 to +14 among treated patients | Proportion | High |

The ten analytic tasks, ordered by increasing complexity and posed identically to the coding agent and the analysts. The Question column summarizes what each task asked (full task prompts in Supplement B), Output the type of result returned, and Complexity our a priori rating. The short name in the Task column identifies each task in Tables 2 to 4.

Two expert SQL analysts (WH and GP) independently wrote queries and answers for each task from these definitions, fixed before testing, then reconciled by adjudication to form the reference standard, which two pediatric infectious diseases physicians (NPM and HTS) reviewed for clinical plausibility. Neither arm saw the other’s work: the analysts built the reference standard blind to the coding agent’s queries and answers, and the agent had no access to the reference standard. In constructing it, the analysts made the institution-specific choices that the schema does not encode, notably excluding visit types recorded as telehealth or virtual even when a face-to-face flag was set, and ascertaining tympanostomy procedures from both the procedure and surgical-procedure sources (Supplement C). Because automated language-model metrics correlate poorly with expert judgment,[17] this adjudicated human standard was the comparator.

JF configured the pipeline. Before the main study, we ran a calibration phase on three questions unrelated to but structured like the study tasks, to verify operation and finalize configuration. We compared three instruction-file lengths (concise, moderate, and verbose); the concise file performed as well as the longer versions and was used throughout (Supplement A). We also attempted two access designs. The design most common in the literature, a mediated layer using the Model Context Protocol (Microsoft Data API Builder), exposed a curated subset of tables one at a time, so the agent could not join across the tables that most questions require; it was slow (about 10 to 15 minutes per question) and often could not complete a query. We therefore gave the agent direct, read-only SQL access over the entire warehouse with iterative execution, under which the calibration questions completed in about 1 to 2 minutes; this design was used throughout.

The index method was a general-purpose autonomous software-engineering agent (OpenAI Codex; model GPT-5.6-sol). It issued queries through a read-only wrapper around a command-line SQL client, run under a dedicated read-only service account with concealed credentials. It read the EHR data dictionary embedded in the warehouse to learn the schema; the application source code was not exposed. Its instruction file (Supplement A) directed it to use the wrapper, run only read-only SQL, use only the codes and parameters provided, and report both the answer and the query. External tools including web access were disabled, and time and token use were logged.

Figure 1 summarizes the design. We evaluated four conditions. First, an autonomous baseline, in which the agent answered each task with no interaction, run three times at medium and three times at extra-high reasoning effort (60 runs). Because extra-high effort produced no accuracy gain over medium (Results), the remaining three conditions used medium effort only. Second, an assumption-listing variant, identical but instructing the agent to list its assumptions before finalizing, three runs per task (30 runs). Third, an interactive session, in which an analyst reviewed the agent’s query against the task definition and gave up to five rounds of bounded feedback. The analyst pointed out in plain language where the query departed from the definition, and could name a field if a hint did not suffice, but never stated the target answer or wrote a query. Each session ended with the agent restating the definition it used (Supplement F). We applied this to four tasks, one session each: the cohort, a procedure count (recurrent-AOM tympanostomy tubes), the time to first tube, and treatment failure. We chose four because live sessions require a human in real time, and these exercise the definitional and case-ascertainment problems behind most baseline errors. Fourth, we tested whether a definition corrected through analyst feedback in the third condition could be reused rather than re-derived for each question. This had two parts. The propagation batch asked whether fixing one definition helps every dependent question. Most tasks build on the same AOM cohort, so we prepended the corrected cohort definition to the five that depend on it (Tasks 3, 4, 5, 6, and 9) and re-ran each three times (15 runs). The consistency batch asked whether a fixed definition gives the same answer on every run. We prepended each interacted task’s own corrected definition and re-ran it five times (20 runs).

**Figure 1.**
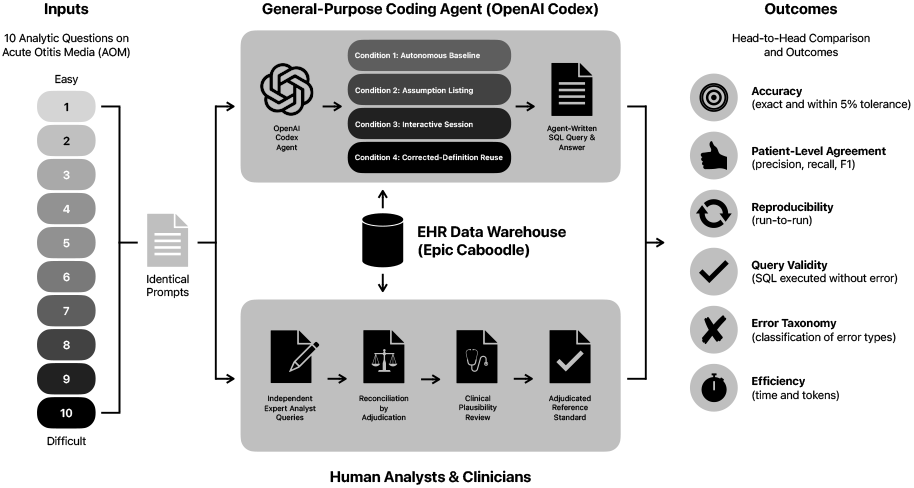
Study design. Identical task prompts (ten analytic tasks) were given to the autonomous coding agent and to expert SQL analysts. The agent wrote and executed its own queries through a read-only interface (dedicated service account, concealed credentials) within a secure, HIPAA-compliant environment; the analysts, working independently and reconciling by adjudication, produced the reference standard, which was also reviewed for clinical plausibility by two pediatric infectious diseases physicians. The agent answered under four conditions (autonomous baseline, assumption listing, interactive session, and corrected-definition reuse). Both queried a full copy of the production data warehouse. Agent answers and the reference standard were compared head-to-head on accuracy, patient-level agreement, reproducibility, query validity, error taxonomy, and efficiency (time and tokens).

We scored each run for accuracy two ways: exact agreement with the reference, meaning the primary value and every breakdown category matched, and agreement within 5% of the primary value. For cohort tasks, we also compared patient sets directly, computing precision, recall, and F1 within the secure environment. Other outcomes were query validity, run-to-run reproducibility, an error taxonomy built by comparing each agent query with the reference query, and efficiency (time and tokens). Analyses were descriptive and summarized at the task level, with the three runs treated as repeated measures rather than independent samples.

## 3 Results

In the first condition (autonomous baseline), all 60 runs produced valid, executable queries, and none fabricated data. Each task was run three times at each effort setting. At medium effort (30 runs), the primary value fell within 5% of the reference on 27 runs and matched it exactly on 10; at extra-high effort it matched exactly on 9 of 30. On the simplest task, counting unique patients, one extra-high run returned 2,148,892 instead of the reference 593,073, because it counted every historical version of each patient’s record rather than each patient once (Table 2).

**Table 2.** Agreement with the Reference Standard (Condition 1: Autonomous Baseline + Condition 3: Interactive Session).

| # | Task | Reference Answer | Exact,<br>Extra-High<br>(/3) | Exact,<br>Medium<br>(/3) | Within<br>5%,<br>Medium<br>(/3) | Interactive<br>Result |
| --- | --- | --- | --- | --- | --- | --- |
| 1 | Birth Month | 593,073 | 0 | 0 | 3 | Not Tested |
| 2 | AOM Cohort | 11,091 | 1 | 1 | 3 | Exact (0 rounds) |
| 3 | Recurrent AOM | 495 | 2 | 0 | 3 | Not Tested |
| 4 | Prescribed Antibiotic | 10,623 of 11,091 | 1 | 1 | 3 | Not Tested |
| 5 | Antibiotic Frequency | See Supplement D | 1 | 3 | 3 | Not Tested |
| 6 | Antibiotic Spectrum Classification | See Supplement D | 2 | 1 | 3 | Not Tested |
| 7 | Tubes (Recurrent) | 55 of 495 | 0 | 0 | 3 | Miss (5 rounds) |
| 8 | Median Days to Tube | 208 days (n = 237) | 1 | 0 | 0 | Within 5% (5 rounds) |
| 9 | Complications | See Supplement D | 1 | 2 | 3 | Not Tested |
| 10 | Treatment Failure | 446 of 10,623 (4.20%) | 0 | 2 | 3 | Exact (2 rounds) |
| **Total** |  |  | **9/30** | **10/30** | **27/30** | **Exact 2 of 4** |

Each task was run three times at each reasoning-effort setting; cells under “Exact” and “Within 5%” give the number of those three runs meeting the criterion. “Exact” means the primary value and every breakdown category matched the adjudicated reference; “within 5%” means the primary value fell within 5% of the reference (the reported count for cohort tasks, the median days for Task 8). The interactive column reports the outcome for the four tasks run with an analyst, with the number of feedback rounds in parentheses (capped at five); the definition-reuse batches (propagation and consistency) are in Supplement G.

Across the three medium-effort runs, the agent gave the same answer on only 3 of the 10 tasks, and only one of those three was also correct (Table 2).

Repeated runs disagreed mainly on one choice, the AOM cohort definition, the most common error class (Table 3). On the same prompt the reported denominator alternated between 11,091, which excludes telehealth and video visits and matches the reference, and 11,114, which includes them, and this single choice propagated into every dependent task (Tasks 3, 4, 5, 6, and 9). The variation was definitional rather than arithmetic; within a run the agent was self-consistent but drew the cohort boundary differently from run to run. In all, 41 of the 60 runs deviated from the reference in at least one value.

**Table 3.** Error Taxonomy.

| Error Class | n | Description and Example |
| --- | --- | --- |
| Cohort-Definition /<br>Denominator Inconsistency | 20 | The agent drew the base AOM cohort differently from run to run, usually by including visit types the definition excluded. On the same prompt it counted telehealth and video visits (11,114) on some runs but excluded them on others (11,091, the reference), and this choice propagated to every dependent task. |
| Valid Query, Incorrect<br>Interpretation | 8 | The SQL executed without error but implemented a different definition than intended. Examples include a different rule for what counts as a treatment-failure antibiotic change (453 vs 446, Task 10) and a miscount of one drug (Task 5). |
| Temporal / Date-Window<br>Error | 6 | The agent applied a different time boundary than the definition, such as a different anchor or inclusivity for the recurrence window. This made the recurrent-AOM cohort range from 469 to 501 across runs instead of the reference 495 (Task 7). |
| Schema-Linking Error | 6 | The agent read from the wrong or an incomplete set of tables. It ascertained tubes from only one of the two procedure sources (228 vs 237, Task 8) and, in the Task 1 outlier, counted historical versions of each patient’s record rather than the current one. |
| Aggregation or Unit Error | 1 | The agent aggregated at the wrong grain, so a duplicated row shifted the count. One duplicate inflated the antibiotic-treated cohort by a single patient (10,624 vs 10,623, Task 4). |
| Missing-Data Handling /<br>Fabrication | 0 | None observed; no run invented values or returned nulls as data. |
| Syntax or Execution Error | 0 | None observed; all 60 runs produced valid, executable SQL. |

Derived from query-by-query comparison of agent and reference queries across all 60 autonomous-baseline runs. Of the 60, 19 matched the reference exactly and 41 deviated in at least one reported value; each deviating run is assigned its primary error class, ordered by frequency.

The agent was fast. The median medium-effort run took 238 seconds and about 499,000 tokens, and the extra-high run 340 seconds and about 616,000 tokens, 2.6 times the reasoning tokens for no accuracy gain.

In the second condition (assumption listing, 30 runs), requiring the agent to state its assumptions did not change accuracy or reproducibility, but the stated assumptions predicted the answer, as runs that named video visits as excluded produced the reference cohort (11,091) and those that did not produced the inflated one (11,114).

In the third condition (interactive session), the agent reached an exact match on two of the four tasks (Supplement E; verbatim feedback in Supplement F). It matched the cohort (Task 2) with no help, and it matched treatment failure (Task 10) after two rounds, once the analyst noted that antibiotics given by feeding tube are enteral routes that still count as systemic therapy, which raised the treated denominator to the reference 10,623. On time to first tube (Task 8), after the analyst pointed it to the surgical-procedure source, the agent recovered the correct patient set (n = 237); its median of 209 days was within 5% of the reference 208 but not exact, a one-day difference in date computation. On the recurrent-tube task (Task 7), it counted tube patients from billing-charge records instead of the procedure sources, overcounting to 97 against the reference 55 (42 spurious patients), and this persisted through all five feedback rounds without reaching the reference.

Finally, in the fourth condition (definition reuse), we tested whether a corrected definition could be reused rather than re-derived. In the propagation batch, with the corrected cohort definition supplied, re-running the five dependent tasks (Tasks 3, 4, 5, 6, and 9) held the denominator at 11,091 in all 15 runs, and 13 of the 15 matched the reference exactly (the two misses were separate task-specific errors). In the consistency batch, we re-ran each interacted task five times with its own corrected definition. Every task returned the same answer on all five runs. The cohort and treatment-failure tasks were correct on all five, whereas the recurrent-tube task repeated its wrong value on all five, because its supplied definition still counted billing charges as tubes (Supplement G).

We then checked whether matching counts reflected the same patients (Table 4). On the six set tasks where the count matched, the patient sets were identical (precision, recall, and F1 all 1.00); the time-to-tube cohort of 237 also matched exactly, localizing its one-day median difference to date computation rather than membership. The exception was the recurrent-tube numerator, where the agent captured all 55 reference tube patients (recall = 1.00) but added 42 from billing charges (precision = 0.57; F1 = 0.72). On treatment failure, agreement was near-complete (445 of 446; F1 = 0.998).

**Table 4.** Patient-Level Agreement with the Reference Standard.

| # | Task | Agent Set | Reference Set | Overlap | Precision | Recall | F1 |
| --- | --- | --- | --- | --- | --- | --- | --- |
| 2 | AOM Cohort | 11,091 | 11,091 | 11,091 | 1.00 | 1.00 | 1.000 |
| 3 | Recurrent AOM | 495 | 495 | 495 | 1.00 | 1.00 | 1.000 |
| 4 | Prescribed Antibiotic | 10,623 | 10,623 | 10,623 | 1.00 | 1.00 | 1.000 |
| 7 | Recurrent Cohort (Denominator) | 495 | 495 | 495 | 1.00 | 1.00 | 1.000 |
| 7 | Tubes (Numerator) | 97 | 55 | 55 | 0.57 | 1.00 | 0.724 |
| 8 | Tube Cohort (Time to First Tube) | 237 | 237 | 237 | 1.00 | 1.00 | 1.000 |
| 9 | Any Complication | 1,253 | 1,253 | 1,253 | 1.00 | 1.00 | 1.000 |
| 10 | Treatment Failure | 446 | 446 | 445 | 1.00 | 1.00 | 0.998 |

Set overlap for each task against the finalized reference standard; F1 is the harmonic mean of precision and recall. The overlaps were computed on a later warehouse snapshot and are accurate to within one patient for the recurrent-tube numerator (Task 7) and treatment failure (Task 10), which does not affect precision or the conclusions. The whole-database birth-month task (Task 1) is omitted because it is not a clinical cohort. Tasks 5 and 6 are omitted because they classify the Task 4 treated cohort, whose patient set matched the reference exactly (F1 = 1.00) and whose category counts matched the reference, so a category-level overlap would add little.

## 4 Discussion

Given open-ended, read-only access to a full copy of a production data warehouse, a general-purpose coding agent answered most routine questions to within a few percent of the adjudicated analyst reference, each within minutes. Working autonomously, however, it matched the reference exactly on fewer than a third of runs and often gave a different answer across its three runs. Reassuringly, the agent never fabricated data, and where its count matched the reference, the patient sets were identical (F1 = 1.00); the exception was the recurrent-tube count, inflated by billing charges. The inconsistency traced to how the agent resolved underspecified definitions, and it did not recur once a definition was fixed.

Reproducibility, not average accuracy, was the main limitation. A query can run without error yet compute the wrong quantity, so a valid query is not a correct answer. On the same prompt, the agent drew the cohort boundary in more than one way, whereas the analyst produced a single value; an answer that shifts between runs cannot be acted on until someone determines which run to trust, and higher reasoning effort did not resolve this. These findings are consistent with evidence that model-written SQL on realistic schemas trails expert performance[2, 3] and that model-drafted clinical logic still requires expert review.[4]

Analyst oversight improved consistency, but only in part. Bounded feedback fixed the definitional errors, and reusing a clarified definition made the dependent tasks reproducible. It did not fix the recurrent-tube count, which stayed wrong through five rounds because the agent kept counting billing charges as tubes. The agent reproduced a wrong definition as faithfully as a right one, so consistency did not guarantee correctness. The correct definition still had to come from a human, because the choices that mattered turned on how the warehouse is set up rather than anything in the data dictionary, such as visit types flagged face-to-face despite being virtual. These details must be supplied in the prompt or as feedback.

How much of the warehouse the agent could see also shaped performance. In calibration, the constrained, curated-subset access that the literature treats as best practice[5, 6, 9] was the main obstacle, and the agent improved once it could query the whole warehouse and its embedded data dictionary. In our judgment, this did not compromise safety. Under both designs the agent had read-only access through a dedicated service account, no visible credentials, no outside tools, and no route to clinical systems, within our HIPAA-compliant environment. These controls applied equally to the curated-subset design. That design therefore added the appearance of tighter security without added protection, while withholding schema context and slowing queries, which degraded its answers. This also sets the evaluation apart from prior clinical agents confined to public research databases, simulated environments, or fixed retrieval tools.[7–10, 13]

This evaluation has limits. It is a single-center feasibility study in one exemplar condition with ten tasks and does not establish generalization across institutions or domains; the reference standard rests on analyst adjudication and clinician plausibility review, not formal validation. We also kept the interactive feedback deliberately general, to avoid biasing the agent and our assessment; more specific guidance might have carried the remaining procedure task to the reference, so the interactive results are a lower bound on what supervised iteration can achieve. Triplicate runs give only a preliminary estimate of reliability, model behavior can drift with version, and because the schema and definitions are not public, training-data contamination is less likely though not excluded. Finally, we evaluated one general-purpose agent at a single version; other agents, or newer versions, might perform differently.

A general-purpose coding agent, given read-only access to the production EHR warehouse, matched an adjudicated analyst reference on most routine questions and returned each answer in minutes. Because its errors were definitional rather than computational, an expert analyst can resolve them by supplying their knowledge of how the warehouse is set up, in the prompt or as feedback. Under that supervision, the agent is already a capable drafting aid that could relieve the analyst backlog constraining research, quality improvement, and operations, and a promising step toward broader hospital analytic support.

## Supporting information

Supplement A. Agent Instruction File (AGENTS.md)

Supplement B. Task Prompts

Supplement C. Analyst Reference SQL Queries

Supplement D. Reference Standard Answers

Supplement E. Interactive Session Summar

Supplement F. Analyst Feedback and Restated Definitions

Supplement G. Definition Reuse (Propagation and Consistency Batches)

Supplement H. SQUIRE 2.0 Checklist

Supplement I. TRIPOD-LLM Checklist

Supplement J. STARD 2015 Checklist

## Statements

### Author contributions

NPM conceived and designed the study, developed the methodology, curated and analyzed the data, and drafted the manuscript. KM originated the project, oversaw it throughout, and provided principal-investigator supervision and project administration. WH served as a reference-standard analyst, wrote the reference queries, and conducted the interactive-session review. GP served as the second reference-standard analyst, independently writing the reference queries. JF built and configured the technical environment, including the Model Context Protocol layer, the read-only SQL wrapper, and the coding-agent and data-warehouse setup, and produced the analysis outputs, including the patient-level comparison. HTS co-developed the ten task questions with NPM and reviewed the results for clinical plausibility. FNH and JHC provided technical expertise in a supervisory role. All authors reviewed and revised the manuscript and approved the final version for submission.

## Acknowledgments

The authors thank OpenAI for technical support in configuring the coding agent during this study and for their engagement in discussions about the study setup.

## Ethics

The activity was determined to be quality improvement and not human subjects research by the Stanford institutional review board (Protocol #88354), with Chief Quality Officer approval; the coding agent was reviewed and approved by the institutional Artificial Intelligence Oversight Committee under a business associate agreement and privacy review.

## Registration

Not registered; this was a quality-improvement activity.

## Patient and public involvement

None.

## Data availability

The underlying data warehouse contains protected health information and cannot be shared. All aggregate results are reported in the manuscript and its supplements.

## Code availability

The agent instruction file (Supplement A), the ten task prompts (Supplement B), and the analyst reference queries with execution results (Supplement C) are provided in full. The warehouse schema and the coding agent are proprietary third-party products and cannot be redistributed.

## Use of AI-assisted technologies

The coding agent evaluated in this study (OpenAI Codex, model GPT-5.6-sol) is the object of the investigation and is described in the Methods; it did not contribute to writing this manuscript. During manuscript preparation, the authors used a large language model assistant to condense and copy-edit author-drafted text and to help organize the tables and supplementary materials. The authors directed all content, verified every number, statement, and citation against primary sources, and take full responsibility for the accuracy, integrity, and originality of the work. No AI tool met authorship criteria or is listed as an author, and no AI-generated text or image was used as source data or cited as a primary source.

## References

1. Everson J, Nong P, and Richwine C. Uptake of generative artificial intelligence integrated with electronic health records in US hospitals. JAMA Netw Open 2025;8:e2549463.

2. Li J, Hui B, Qu G, Yang J, Li B, Li B, et al. Can LLM already serve as a database interface? A big bench for large-scale database grounded text-to-SQLs (BIRD). In: Advances in Neural Information Processing Systems. Vol. 36. arXiv:2305.03111. 2023.

3. Lei F, Chen J, Ye Y, Cao R, Shin D, Su H, et al. Spider 2.0: evaluating language models on real-world enterprise text-to-SQL workflows. In: International Conference on Learning Representations (ICLR). arXiv:2411.07763. 2025.

4. Yan C, Ong HH, Grabowska ME, Krantz MS, Su WC, Dickson AL, et al. Large language models facilitate the generation of electronic health record phenotyping algorithms. J Am Med Inform Assoc 2024;31:1994–2001.

5. Gallifant J, Kellogg KC, Butler M, Centi A, Chen S, Doyle P, et al. Beyond the algorithm: a field guide to deploying AI agents in clinical practice. arXiv:2509.26153. Preprint. 2025.

6. Xu G, Li X, Chen Y, Duan Y, Wu S, Yu H, et al. A comprehensive survey of AI agents in healthcare. J Biomed Inform 2026;179:105045.

7. Shi W, Xu R, Zhuang Y, Yu Y, Zhang J, Wu H, et al. EHRAgent: code empowers large language models for few-shot complex tabular reasoning on electronic health records. In: Proceedings of the 2024 Conference on Empirical Methods in Natural Language Processing (EMNLP). 2024:22315–39. doi: 10.18653/v1/2024.emnlp-main.1245.

8. Lee K, Hong S, Park J, Lim J, Choi J, Yoon D, et al. EMR-AGENT: automating cohort and feature extraction from EMR databases. arXiv:2510.00549. Preprint. 2025.

9. Masayoshi K, Hashimoto M, Yokoyama R, Toda N, Uwamino Y, Fukuda S, et al. EHR-MCP: real-world evaluation of clinical information retrieval by large language models via Model Context Protocol. arXiv:2509.15957. Preprint. 2025.

10. Jiang Y, Black KC, Geng G, Park D, Zou J, Ng AY, et al. MedAgentBench: a virtual EHR environment to benchmark medical LLM agents. NEJM AI 2025;2.

11. Shah NH, Ambers N, Pandya A, Keyes T, Banda JM, Nallan S, et al. Adoption and use of LLMs at an academic medical center. arXiv:2602.00074. Preprint. 2026.

12. Low YS, Jackson ML, Hyde RJ, Brown RE, Sanghavi NM, Baldwin JD, et al. Answering real-world clinical questions using large language model, retrieval-augmented generation, and agentic systems. Digit Health 2025;11:20552076251348850.

13. Ferber D, Hilgers L, Höper C, Kinny-Köster B, Eckardt JN, Egger-Heidrich K, et al. Towards autonomous medical artificial intelligence agents. Nature 2026.

14. Ogrinc G, Davies L, Goodman D, Batalden P, Davidoff F, and Stevens D. SQUIRE 2.0 (Standards for Quality Improvement Reporting Excellence): revised publication guidelines from a detailed consensus process. BMJ Qual Saf 2016;25:986–92.

15. Gallifant J, Afshar M, Ameen S, Aphinyanaphongs Y, Chen S, Cacciamani G, et al. The TRIPOD-LLM reporting guideline for studies using large language models. Nat Med 2025;31:60–9.

16. Bossuyt PM, Reitsma JB, Bruns DE, Gatsonis CA, Glasziou PP, Irwig L, et al. STARD 2015: an updated list of essential items for reporting diagnostic accuracy studies. BMJ 2015;351:h5527.

17. Du X, Zhou Z, Wang Y, Chuang YW, Li Y, Yang R, et al. Testing and evaluation of generative large language models in electronic health record applications: a systematic review. J Am Med Inform Assoc 2026;33:743–53.

