## Supplement A. Agent Instruction File (AGENTS.md) for "Can a General-Purpose Coding Agent Analyze a Production Hospital Data Warehouse?"

```
Use `study\query-cdw.ps1` (sqlcmd wrapper) for all database queries.

Prefer the "FullAccess" database schema for all queries.  (Not for schema lookups)

Invoke it as:
& '.\study\query-cdw.ps1' -Query '<SQL>'

Do not open, read, display, modify, or summarize the wrapper or its connection
string. Never print credentials or environment variables. Execute read-only SQL
only.

Derive every answer solely from your own SQL queries against the database. Do
not read, list, or grep any other file on disk; use only the query wrapper.

You have read-only access to an Epic Caboodle EHR data warehouse. Answer the
task below. Use only the codes and parameters given. Report the final answer
and the SQL you used.
```

*The instruction file used in production named the wrapper by its local path; that path is genericized here. The line directing the agent to derive answers only from its own SQL and not to read other files on disk prevented it from inspecting the task prompts, the reference definitions, or other runs.*
