## Supplement B. Task Prompts for "Can a General-Purpose Coding Agent Analyze a Production Hospital Data Warehouse?"

### Task 1. Birth Month

- **Question:** How many unique patients in the database have a date of birth in September? Count each patient once (one row per unique patient). The parameters are specified below.
- **Population:** all patients in the database (no diagnosis or setting restriction).
- **Report:** the count, and the SQL you used.

### Task 2. AOM Cohort

- **Question:** How many unique patients were diagnosed with acute otitis media (AOM) at an SMCH primary care pediatric outpatient clinic during the index window? Count each patient once. This defines the AOM cohort used by later tasks. The definition is specified below.
- **AOM cohort definition (use this exact definition in every task that references the AOM cohort):**
  - patients younger than 26 years at the encounter date;
  - with an AOM diagnosis (ICD-10 H66.001 to H66.019, H66.40 to H66.43, H66.90 to H66.93) **recorded as an encounter diagnosis** (not a problem-list or historical diagnosis);
  - at a **completed, in-person (face-to-face) outpatient** encounter;
  - at an SMCH primary care pediatric outpatient clinic: department_id in (10801100, 10802100, 10803100, 10804100, 10806100, 10807100, 10808100, 10809100, 10810100, 10814100, 10815100, 10816100, 10829100, 10830100, 10837100, 10838100, 10839100, 10842100, 10843100, 10847100, 10850105, 10855101, 10860100, 10861100, 10863100, 10864100, 10865102, 10867100, 10868001, 10873100);
  - during 2023-01-01 through 2024-12-31.
- **Question restated:** How many unique patients meet this definition? Count each patient once.
- **Report:** the count, and the SQL you used.

### Task 3. Recurrent AOM

- **Question:** Of the AOM cohort (defined below), how many patients meet the definition of recurrent AOM (defined below)? The cohort, look-back, and recurrence definition are specified below.
- **AOM cohort:** patients younger than 26 at the encounter date with an AOM diagnosis (ICD-10 H66.001 to H66.019, H66.40 to H66.43, H66.90 to H66.93) recorded as an encounter diagnosis at a completed, in-person outpatient visit at an SMCH primary care pediatric outpatient clinic (department_id in (10801100, 10802100, 10803100, 10804100, 10806100, 10807100, 10808100, 10809100, 10810100, 10814100, 10815100, 10816100, 10829100, 10830100, 10837100, 10838100, 10839100, 10842100, 10843100, 10847100, 10850105, 10855101, 10860100, 10861100, 10863100, 10864100, 10865102, 10867100, 10868001, 10873100)) during 2023-01-01 through 2024-12-31.
- **Look-back:** include AOM encounters from 2022-01-01 onward when identifying episodes.
- **Recurrence definition (precise):** a patient is recurrent if there exist three AOM encounters for that patient such that the second is at least 30 days after the first, the third is at least 30 days after

the second, and all three fall within a single 6-month window (third encounter no later than 6 months after the first). Same-day or <30-day-apart encounters do not count as separate episodes.

- **Question restated:** How many AOM cohort patients meet this recurrence definition?
- **Report:** the numerator (recurrent patients), the denominator (AOM cohort), and the SQL you used.

### Task 4. Prescribed Antibiotic

- **Question:** Of the AOM cohort (defined below), how many patients were prescribed a systemic antibiotic within 3 days of the AOM diagnosis? The cohort, antibiotic definition, and linkage are specified below.
- **AOM cohort:** patients younger than 26 at the encounter date with an AOM diagnosis (ICD-10 H66.001 to H66.019, H66.40 to H66.43, H66.90 to H66.93) recorded as an encounter diagnosis at a completed, in-person outpatient visit at an SMCH primary care pediatric outpatient clinic (department_id in (10801100, 10802100, 10803100, 10804100, 10806100, 10807100, 10808100, 10809100, 10810100, 10814100, 10815100, 10816100, 10829100, 10830100, 10837100, 10838100, 10839100, 10842100, 10843100, 10847100, 10850105, 10855101, 10860100, 10861100, 10863100, 10864100, 10865102, 10867100, 10868001, 10873100)) during 2023-01-01 through 2024-12-31.
- **Antibiotic definition:** a systemic (oral or parenteral) antibacterial, identified by its therapeutic/pharmaceutical class or by ingredient name; exclude otic, ophthalmic, and topical routes.
- **Linkage and location:** the antibiotic must be ordered at one of these clinics, on the AOM diagnosis date through 3 days after (Day 0 to Day 3).
- **Question restated:** How many AOM cohort patients had such an antibiotic?
- **Report:** the numerator, the denominator (AOM cohort), and the SQL you used.

### Task 5. Antibiotic Frequency

- **Question:** Among the antibiotic-treated AOM cohort (defined below), report the number and percentage of patients receiving each of the five named antibiotics, with all other systemic antibacterials grouped as "other." The cohort, named antibiotics, and counting rule are specified below.
- **Antibiotic-treated AOM cohort:** patients younger than 26 at the encounter date with an AOM diagnosis (ICD-10 H66.001 to H66.019, H66.40 to H66.43, H66.90 to H66.93) recorded as an encounter diagnosis at a completed, in-person outpatient visit at an SMCH primary care pediatric outpatient clinic (department_id in (10801100, 10802100, 10803100, 10804100, 10806100, 10807100, 10808100, 10809100, 10810100, 10814100, 10815100, 10816100, 10829100, 10830100, 10837100, 10838100, 10839100, 10842100, 10843100, 10847100, 10850105, 10855101, 10860100, 10861100, 10863100, 10864100, 10865102, 10867100, 10868001, 10873100)) during 2023-01-01 through 2024-12-31, who were prescribed a systemic (oral or parenteral) antibacterial at one of these clinics within 3 days of the AOM diagnosis (Day 0 to Day 3).
- **Named antibiotics:**
  - amoxicillin (exclude amoxicillin-clavulanate)
  - amoxicillin-clavulanate
  - cefdinir
  - clindamycin
  - ceftriaxone

  - other (all other systemic antibacterials)

- **Counting rule:** count all qualifying antibiotics in the 3-day window. A patient can appear in more than one category, so category percentages may total more than 100%.
- **Report:** counts and percentages by category, the denominator (antibiotic-treated cohort), and the SQL you used.

### Task 6. Antibiotic Spectrum Classification

- **Question:** Among the antibiotic-treated AOM cohort (defined below), what proportion of patients received a narrow-spectrum, broad-spectrum, or other antibiotic as their FIRST AOM antibiotic? The cohort and spectrum definitions are specified below.
- **Antibiotic-treated AOM cohort:** patients younger than 26 at the encounter date with an AOM diagnosis (ICD-10 H66.001 to H66.019, H66.40 to H66.43, H66.90 to H66.93) recorded as an encounter diagnosis at a completed, in-person outpatient visit at an SMCH primary care pediatric outpatient clinic (department_id in (10801100, 10802100, 10803100, 10804100, 10806100, 10807100, 10808100, 10809100, 10810100, 10814100, 10815100, 10816100, 10829100, 10830100, 10837100, 10838100, 10839100, 10842100, 10843100, 10847100, 10850105, 10855101, 10860100, 10861100, 10863100, 10864100, 10865102, 10867100, 10868001, 10873100)) during 2023-01-01 through 2024-12-31, who were prescribed a systemic (oral or parenteral) antibacterial at one of these clinics within 3 days of the AOM diagnosis (Day 0 to Day 3).
- **First antibiotic:** the earliest qualifying antibiotic by order time; break ties deterministically. Each patient is classified into exactly one category, so the three categories sum to 100%.
- **Spectrum definitions:**

  - **narrow:** amoxicillin (excluding amoxicillin-clavulanate)
  - **broad:** amoxicillin-clavulanate, cefdinir, clindamycin, or ceftriaxone
  - **other:** any systemic antibacterial not in the narrow or broad sets

- **Report:** counts and percentages by category, the denominator (antibiotic-treated cohort), and the SQL you used.

### Task 7. Tubes (Recurrent)

- **Question:** Among the recurrent AOM cohort (defined below), how many patients underwent tympanostomy tube placement during the study window? The cohort, procedure codes, ascertainment, and dates are specified below.
- **Recurrent AOM cohort:** AOM cohort patients (patients younger than 26 at the encounter date with an AOM diagnosis, ICD-10 H66.001 to H66.019 / H66.40 to H66.43 / H66.90 to H66.93, recorded as an encounter diagnosis at a completed, in-person outpatient visit at an SMCH primary care pediatric outpatient clinic (department_id in (10801100, 10802100, 10803100, 10804100, 10806100, 10807100, 10808100, 10809100, 10810100, 10814100, 10815100, 10816100, 10829100, 10830100, 10837100, 10838100, 10839100, 10842100, 10843100, 10847100, 10850105, 10855101, 10860100, 10861100, 10863100, 10864100, 10865102, 10867100, 10868001, 10873100)) during 2023-01-01 through 2024-12-31) who meet the recurrence definition: three AOM encounters, each at least 30 days after the previous, all within a 6-month window, using look-back to 2022-01-01.

- **Tympanostomy tube codes (CPT):** 69433, 69436, 0583T. Exclude 69420 and 69421 (myringotomy without tube) and 69424 (tube removal).
- **Ascertainment and window:** ascertain tube procedures system-wide (not limited to these clinics). Count a tube only if it occurs on or after the patient's first AOM diagnosis (the earliest AOM encounter in the index window, 2023-2024), through 2025-12-31. Do not apply a fixed 2023-01-01 start cutoff independent of the patient's first AOM.
- **Report:** the numerator (patients with a tube), the denominator (recurrent cohort), and the SQL you used.

### Task 8. Median Days to Tube

- **Question:** Among AOM cohort patients who underwent tympanostomy tube placement on or after their first AOM diagnosis, what is the median and interquartile range of the number of days from the first AOM diagnosis to the first such tube? The cohort, procedure codes, and dates are specified below.
- **Cohort:** AOM cohort patients (patients younger than 26 at the encounter date with an AOM diagnosis, ICD-10 H66.001 to H66.019 / H66.40 to H66.43 / H66.90 to H66.93, recorded as an encounter diagnosis at a completed, in-person outpatient visit at an SMCH primary care pediatric outpatient clinic (department_id in (10801100, 10802100, 10803100, 10804100, 10806100, 10807100, 10808100, 10809100, 10810100, 10814100, 10815100, 10816100, 10829100, 10830100, 10837100, 10838100, 10839100, 10842100, 10843100, 10847100, 10850105, 10855101, 10860100, 10861100, 10863100, 10864100, 10865102, 10867100, 10868001, 10873100)) during 2023-01-01 through 2024-12-31).
- **Tube:** the first tympanostomy (CPT 69433, 69436, or 0583T; ascertain system-wide) that occurs ON OR AFTER the patient's first AOM diagnosis, through 2025-12-31. Do not count tubes placed before the first AOM diagnosis.
- **Question restated:** For patients with such a tube, compute days from first AOM diagnosis to first tube; report median, interquartile range, and the number of patients included.
- **Report:** the median, the interquartile range, the number of patients, and the SQL you used.

### Task 9. Complications

- **Question:** How many AOM cohort patients developed any of the specified complications? Report the total with any complication and a breakdown by type. The cohort and complication definitions are specified below.
- **AOM cohort:** patients younger than 26 at the encounter date with an AOM diagnosis (ICD-10 H66.001 to H66.019, H66.40 to H66.43, H66.90 to H66.93) recorded as an encounter diagnosis at a completed, in-person outpatient visit at an SMCH primary care pediatric outpatient clinic (department_id in (10801100, 10802100, 10803100, 10804100, 10806100, 10807100, 10808100, 10809100, 10810100, 10814100, 10815100, 10816100, 10829100, 10830100, 10837100, 10838100, 10839100, 10842100, 10843100, 10847100, 10850105, 10855101, 10860100, 10861100, 10863100, 10864100, 10865102, 10867100, 10868001, 10873100)) during 2023-01-01 through 2024-12-31.
- **Complication definitions (ascertain system-wide):**
  - **Accompanying conjunctivitis (otitis-conjunctivitis syndrome):** an acute conjunctivitis diagnosis (ICD-10 H10.0 to H10.3) on the same encounter as an AOM diagnosis, or within 7 days after it (Day 0 to Day +7); conjunctivitis coded before the AOM diagnosis does not count.

- **Tympanic membrane perforation:** ICD-10 H72.0 to H72.9, recorded on or after the patient's first AOM diagnosis, through 2025-12-31.
- **Acute mastoiditis:** ICD-10 H70.0 and subcodes, recorded on or after the patient's first AOM diagnosis, through 2025-12-31.

- **Question restated:** How many AOM cohort patients had any of these? Report the total (each patient counted once) and the count for each type.
- **Report:** the total, the breakdown by type, the denominator (AOM cohort), and the SQL you used.

### Task 10. Treatment Failure

- **Question:** Among the antibiotic-treated AOM cohort (defined below), what fraction of patients had treatment failure (defined below)? The cohort and treatment-failure definition are specified below.
- **Antibiotic-treated AOM cohort:** patients younger than 26 at the encounter date with an AOM diagnosis (ICD-10 H66.001 to H66.019, H66.40 to H66.43, H66.90 to H66.93) recorded as an encounter diagnosis at a completed, in-person outpatient visit at an SMCH primary care pediatric outpatient clinic (department_id in (10801100, 10802100, 10803100, 10804100, 10806100, 10807100, 10808100, 10809100, 10810100, 10814100, 10815100, 10816100, 10829100, 10830100, 10837100, 10838100, 10839100, 10842100, 10843100, 10847100, 10850105, 10855101, 10860100, 10861100, 10863100, 10864100, 10865102, 10867100, 10868001, 10873100)) during 2023-01-01 through 2024-12-31, who were prescribed a systemic (oral or parenteral) antibacterial at one of these clinics within 3 days of the AOM diagnosis (Day 0 to Day 3). The index encounter is the AOM encounter of that first antibiotic (Day 0).
- **Treatment failure definition:** a return visit at one of these clinics on Day +2 through Day +14 (relative to the index encounter) at which a different antibiotic agent is ordered. A refill or re-order of the same agent does not count; compare at the ingredient/agent level.
- **Question restated:** What fraction of the antibiotic-treated cohort had treatment failure?
- **Report:** the numerator, the denominator (antibiotic-treated cohort), the percentage, and the SQL you used.
