## Supplement C. Analyst Reference SQL Queries for "Can a General-Purpose Coding Agent Analyze a Production Hospital Data Warehouse?"

The reference standard was implemented as ten task queries, each self-contained: every query inlines the shared central definitions it uses (the qualifying AOM encounter and patient cohort, and, where needed, the recurrence, tympanostomy, and antibiotic definitions), so it can be run on its own. These queries are the analyst-side counterpart to the plain-language task prompts in Supplement B, and each was confirmed against its prompt. Each query is reproduced below exactly as executed, with an execution-result header recording the finalized answer from the 2026-07-31 warehouse refresh; those answers are the reference standard summarized in Supplement D.

### Task 1. Birth Month

```
/*
EXECUTION RESULT
Run date: 2026-07-31 (America/Los_Angeles)
Command: uv run python query.py -f .\queries\01_unique_patients_born_in_month.sql

UniquePatientsBornInMonth: 593073
*/

/*
Task 1: Patients born in a given month

Shared definitions: none. This task uses only dbo.PatientDim.
*/

DECLARE @BirthMonth TINYINT = 9;

SELECT
    COUNT(DISTINCT p.DurableKey) AS UniquePatientsBornInMonth
FROM dbo.PatientDim AS p
WHERE p.IsCurrent = 1
  AND p.BirthDate IS NOT NULL
  AND MONTH(p.BirthDate) = @BirthMonth;
```

### Task 2. AOM Cohort

```
/*
EXECUTION RESULT
Run date: 2026-07-31 (America/Los_Angeles)
Command: uv run python query.py -f .\queries\02_unique_patients_under_26_aom.sql

AomCohortPatients: 11091
*/

/*
Task 2: AOM cohort

Shared definition copied below:
```

```sql
- definitions/AOM-ENCOUNTER.sql
*/

DECLARE @AomStartDate DATE = '2023-01-01';
DECLARE @AomEndDate   DATE = '2024-12-31';

/* BEGIN SHARED DEFINITION: definitions/AOM-ENCOUNTER.sql (AOM-ENC v1.0)
   CTEs: smch_pediatric_departments, aom_diagnosis_keys, aom_encounters */

-- Exact protocol-supplied SMCH primary care pediatric departments.
-- All 30 IDs mapped and had utilization during validation. Keep the explicitly
-- supplied ZZZ-named historical IDs; do not infer an additional active-status
-- restriction from the department name.
WITH smch_pediatric_departments AS (
    SELECT v.DepartmentEpicId
    FROM (VALUES
        ('10801100'), ('10802100'), ('10803100'), ('10804100'), ('10806100'),
        ('10807100'), ('10808100'), ('10809100'), ('10810100'), ('10814100'),
        ('10815100'), ('10816100'), ('10829100'), ('10830100'), ('10837100'),
        ('10838100'), ('10839100'), ('10842100'), ('10843100'), ('10847100'),
        ('10850105'), ('10855101'), ('10860100'), ('10861100'), ('10863100'),
        ('10864100'), ('10865102'), ('10867100'), ('10868001'), ('10873100')
    ) AS v(DepartmentEpicId)
),

-- Exact AOM ICD-10-CM inclusion:
-- H66.001-H66.019, H66.40-H66.43, and H66.90-H66.93.
-- Resolving DiagnosisKey first handles multiple terminology rows per code.
aom_diagnosis_keys AS (
    SELECT DISTINCT
        dt.DiagnosisKey
    FROM dbo.DiagnosisTerminologyDim AS dt
    WHERE dt.Type = 'ICD-10-CM'
      AND (
            dt.Value LIKE 'H66.00[1-9]'
         OR dt.Value LIKE 'H66.01[0-9]'
         OR dt.Value IN (
                'H66.40', 'H66.41', 'H66.42', 'H66.43',
                'H66.90', 'H66.91', 'H66.92', 'H66.93'
         )
      )
),

-- One row per patient and qualifying AOM encounter. Retaining EncounterKey and
-- EncounterDate supports recurrence, antibiotic linkage, and follow-up tasks.
aom_encounters AS (
    SELECT DISTINCT
        def.PatientDurableKey,
        def.EncounterKey,
        CAST(ef.Date AS DATE) AS EncounterDate,
        dd.DepartmentEpicId
    FROM dbo.DiagnosisEventFact AS def
    INNER JOIN aom_diagnosis_keys AS aom
        ON aom.DiagnosisKey = def.DiagnosisKey
```

```sql
    -- Require the diagnosis to be attached to a real encounter.
    INNER JOIN dbo.EncounterFact AS ef
        ON ef.EncounterKey = def.EncounterKey

    -- Apply the supplied clinic list through the encounter's historical
    -- DepartmentKey and stable DepartmentEpicId.
    INNER JOIN dbo.DepartmentDim AS dd
        ON dd.DepartmentKey = ef.DepartmentKey
    INNER JOIN smch_pediatric_departments AS smch
        ON smch.DepartmentEpicId = dd.DepartmentEpicId

    -- PatientDim is history-tracked. Use exactly one current patient record.
    INNER JOIN dbo.PatientDim AS p
        ON p.DurableKey = def.PatientDurableKey
       AND p.IsCurrent = 1

    -- Diagnosis source: encounter diagnosis only. Explicitly exclude billing
    -- diagnoses, medical history, and problem-list diagnoses.
    WHERE def.Type = 'Encounter Diagnosis'

      -- Completed in-person outpatient encounter.
      -- The Caboodle face-to-face flag alone included telehealth/video visits,
      -- so also exclude video flags and virtual/phone VisitType labels.
      AND ef.DerivedEncounterStatus = 'Complete'
      AND ef.IsOutpatientFaceToFaceVisit = 1
      AND COALESCE(ef.IsVideoVisit, 0) = 0
      AND UPPER(COALESCE(ef.VisitType, '')) NOT LIKE '%TELEHEALTH%'
      AND UPPER(COALESCE(ef.VisitType, '')) NOT LIKE '%VIRTUAL%'
      AND UPPER(COALESCE(ef.VisitType, '')) NOT LIKE '%PHONE%'

      -- Inclusive calendar-date window that remains correct when ef.Date
      -- contains a time component.
      AND ef.Date >= @AomStartDate
      AND ef.Date < DATEADD(DAY, 1, @AomEndDate)

      -- Younger than 26 on the encounter date. This direct birthday comparison
      -- excludes future birth dates and excludes patients on/after age 26.
      -- Approved leap-day convention: SQL Server treats February 28 as the
      -- birthday in a non-leap year; no validated patients were affected.
      AND p.BirthDate IS NOT NULL
      AND p.BirthDate <= CAST(ef.Date AS DATE)
      AND CAST(ef.Date AS DATE) < DATEADD(YEAR, 26, p.BirthDate)

      -- Valid analytic patient only. These safeguards changed neither the
      -- validated index cohort nor the look-back population.
      AND p.IsValid = 1
      AND p.Test = 0
      AND p.DurableKey > 0
)

/* END SHARED DEFINITION: definitions/AOM-ENCOUNTER.sql */

/* BEGIN TASK-SPECIFIC LOGIC: Task 2 */
```

```sql
SELECT
    COUNT(DISTINCT ae.PatientDurableKey) AS AomCohortPatients
FROM aom_encounters AS ae;

/* END TASK-SPECIFIC LOGIC: Task 2 */
```

### Task 3. Recurrent AOM

```sql
/*
EXECUTION RESULT
Run date: 2026-07-31 (America/Los_Angeles)
Command: uv run python query.py -f .\queries\03_recurrent_aom_3_episodes_6_months.sql

RecurrentAomPatients: 495
AomCohortPatients:     11091
*/

/*
Task 3: Recurrent AOM

Shared definitions copied below:
- AOM-ENCOUNTER uses the 2022 look-back start for this task.
- AOM-RECURRENT separately limits index-cohort membership to 2023-2024.
*/

DECLARE @LookbackStartDate DATE = '2022-01-01';
DECLARE @StudyStartDate    DATE = '2023-01-01';
DECLARE @StudyEndDate      DATE = '2024-12-31';
DECLARE @AomStartDate      DATE = @LookbackStartDate;
DECLARE @AomEndDate        DATE = @StudyEndDate;

/* BEGIN SHARED DEFINITION: definitions/AOM-ENCOUNTER.sql (AOM-ENC v1.0)
   CTEs: smch_pediatric_departments, aom_diagnosis_keys, aom_encounters */

-- Exact protocol-supplied SMCH primary care pediatric departments.
-- All 30 IDs mapped and had utilization during validation. Keep the explicitly
-- supplied ZZZ-named historical IDs; do not infer an additional active-status
-- restriction from the department name.
WITH smch_pediatric_departments AS (
    SELECT v.DepartmentEpicId
    FROM (VALUES
        ('10801100'), ('10802100'), ('10803100'), ('10804100'), ('10806100'),
        ('10807100'), ('10808100'), ('10809100'), ('10810100'), ('10814100'),
        ('10815100'), ('10816100'), ('10829100'), ('10830100'), ('10837100'),
        ('10838100'), ('10839100'), ('10842100'), ('10843100'), ('10847100'),
        ('10850105'), ('10855101'), ('10860100'), ('10861100'), ('10863100'),
        ('10864100'), ('10865102'), ('10867100'), ('10868001'), ('10873100')
    ) AS v(DepartmentEpicId)
),

-- Exact AOM ICD-10-CM inclusion:
```

```sql
-- H66.001-H66.019, H66.40-H66.43, and H66.90-H66.93.
-- Resolving DiagnosisKey first handles multiple terminology rows per code.
aom_diagnosis_keys AS (
    SELECT DISTINCT
        dt.DiagnosisKey
    FROM dbo.DiagnosisTerminologyDim AS dt
    WHERE dt.Type = 'ICD-10-CM'
      AND (
            dt.Value LIKE 'H66.00[1-9]'
         OR dt.Value LIKE 'H66.01[0-9]'
         OR dt.Value IN (
                'H66.40', 'H66.41', 'H66.42', 'H66.43',
                'H66.90', 'H66.91', 'H66.92', 'H66.93'
         )
      )
),

-- One row per patient and qualifying AOM encounter. Retaining EncounterKey and
-- EncounterDate supports recurrence, antibiotic linkage, and follow-up tasks.
aom_encounters AS (
    SELECT DISTINCT
        def.PatientDurableKey,
        def.EncounterKey,
        CAST(ef.Date AS DATE) AS EncounterDate,
        dd.DepartmentEpicId
    FROM dbo.DiagnosisEventFact AS def
    INNER JOIN aom_diagnosis_keys AS aom
        ON aom.DiagnosisKey = def.DiagnosisKey

    -- Require the diagnosis to be attached to a real encounter.
    INNER JOIN dbo.EncounterFact AS ef
        ON ef.EncounterKey = def.EncounterKey

    -- Apply the supplied clinic list through the encounter's historical
    -- DepartmentKey and stable DepartmentEpicId.
    INNER JOIN dbo.DepartmentDim AS dd
        ON dd.DepartmentKey = ef.DepartmentKey
    INNER JOIN smch_pediatric_departments AS smch
        ON smch.DepartmentEpicId = dd.DepartmentEpicId

    -- PatientDim is history-tracked. Use exactly one current patient record.
    INNER JOIN dbo.PatientDim AS p
        ON p.DurableKey = def.PatientDurableKey
       AND p.IsCurrent = 1

    -- Diagnosis source: encounter diagnosis only. Explicitly exclude billing
    -- diagnoses, medical history, and problem-list diagnoses.
    WHERE def.Type = 'Encounter Diagnosis'

      -- Completed in-person outpatient encounter.
      -- The Caboodle face-to-face flag alone included telehealth/video visits,
      -- so also exclude video flags and virtual/phone VisitType labels.
      AND ef.DerivedEncounterStatus = 'Complete'
      AND ef.IsOutpatientFaceToFaceVisit = 1
```

```sql
      AND COALESCE(ef.IsVideoVisit, 0) = 0
      AND UPPER(COALESCE(ef.VisitType, '')) NOT LIKE '%TELEHEALTH%'
      AND UPPER(COALESCE(ef.VisitType, '')) NOT LIKE '%VIRTUAL%'
      AND UPPER(COALESCE(ef.VisitType, '')) NOT LIKE '%PHONE%'

      -- Inclusive calendar-date window that remains correct when ef.Date
      -- contains a time component.
      AND ef.Date >= @AomStartDate
      AND ef.Date < DATEADD(DAY, 1, @AomEndDate)

      -- Younger than 26 on the encounter date. This direct birthday comparison
      -- excludes future birth dates and excludes patients on/after age 26.
      -- Approved leap-day convention: SQL Server treats February 28 as the
      -- birthday in a non-leap year; no validated patients were affected.
      AND p.BirthDate IS NOT NULL
      AND p.BirthDate <= CAST(ef.Date AS DATE)
      AND CAST(ef.Date AS DATE) < DATEADD(YEAR, 26, p.BirthDate)

      -- Valid analytic patient only. These safeguards changed neither the
      -- validated index cohort nor the look-back population.
      AND p.IsValid = 1
      AND p.Test = 0
      AND p.DurableKey > 0
)

/* END SHARED DEFINITION: definitions/AOM-ENCOUNTER.sql */

/* BEGIN SHARED DEFINITION: definitions/AOM-RECURRENT.sql (AOM-REC v0.2)
   CTEs: aom_index_cohort, aom_episode_dates,
         candidate_recurrence_triplets, recurrent_aom_cohort */

, aom_index_cohort AS (
    SELECT DISTINCT
        ae.PatientDurableKey
    FROM aom_encounters AS ae
    WHERE ae.EncounterDate >= @StudyStartDate
      AND ae.EncounterDate < DATEADD(DAY, 1, @StudyEndDate)
),
aom_episode_dates AS (
    SELECT DISTINCT
        ae.PatientDurableKey,
        ae.EncounterDate AS EpisodeDate
    FROM aom_encounters AS ae
),
candidate_recurrence_triplets AS (
    SELECT
        first_episode.PatientDurableKey,
        first_episode.EpisodeDate AS Episode1Date,
        second_episode.EpisodeDate AS Episode2Date,
        third_episode.EpisodeDate AS Episode3Date
    FROM aom_episode_dates AS first_episode
    INNER JOIN aom_episode_dates AS second_episode
        ON second_episode.PatientDurableKey = first_episode.PatientDurableKey
       AND second_episode.EpisodeDate
```

```sql
            >= DATEADD(DAY, 30, first_episode.EpisodeDate)
    INNER JOIN aom_episode_dates AS third_episode
        ON third_episode.PatientDurableKey = second_episode.PatientDurableKey
       AND third_episode.EpisodeDate
            >= DATEADD(DAY, 30, second_episode.EpisodeDate)
       AND third_episode.EpisodeDate
            <= DATEADD(MONTH, 6, first_episode.EpisodeDate)
),
recurrent_aom_cohort AS (
    SELECT DISTINCT
        cohort.PatientDurableKey
    FROM aom_index_cohort AS cohort
    INNER JOIN candidate_recurrence_triplets AS triplet
        ON triplet.PatientDurableKey = cohort.PatientDurableKey
)

/* END SHARED DEFINITION: definitions/AOM-RECURRENT.sql */

/* BEGIN TASK-SPECIFIC LOGIC: Task 3 */

SELECT
    (SELECT COUNT(*) FROM recurrent_aom_cohort) AS RecurrentAomPatients,
    (SELECT COUNT(*) FROM aom_index_cohort) AS AomCohortPatients;

/* END TASK-SPECIFIC LOGIC: Task 3 */
```

### Task 4. Prescribed Antibiotic

```sql
/*
EXECUTION RESULT
Run date: 2026-07-31 (America/Los_Angeles)
Command: uv run python query.py -f .\queries\04_aom_patients_with_systemic_antibiotic_3_days.sql

PatientsWithSystemicAntibioticWithin3Days: 10623
AomCohortPatients:                          11091
*/

/*
Task 4: Antibiotic prescribed within 3 days

Shared definitions copied below:
- definitions/AOM-ENCOUNTER.sql
- definitions/AOM-ANTIBIOTIC.sql
*/

DECLARE @StudyStartDate DATE = '2023-01-01';
DECLARE @StudyEndDate   DATE = '2024-12-31';
DECLARE @AomStartDate   DATE = @StudyStartDate;
DECLARE @AomEndDate     DATE = @StudyEndDate;

/* BEGIN SHARED DEFINITION: definitions/AOM-ENCOUNTER.sql (AOM-ENC v1.0)
   CTEs: smch_pediatric_departments, aom_diagnosis_keys, aom_encounters */
```

```sql
-- Exact protocol-supplied SMCH primary care pediatric departments.
-- All 30 IDs mapped and had utilization during validation. Keep the explicitly
-- supplied ZZZ-named historical IDs; do not infer an additional active-status
-- restriction from the department name.
WITH smch_pediatric_departments AS (
    SELECT v.DepartmentEpicId
    FROM (VALUES
        ('10801100'), ('10802100'), ('10803100'), ('10804100'), ('10806100'),
        ('10807100'), ('10808100'), ('10809100'), ('10810100'), ('10814100'),
        ('10815100'), ('10816100'), ('10829100'), ('10830100'), ('10837100'),
        ('10838100'), ('10839100'), ('10842100'), ('10843100'), ('10847100'),
        ('10850105'), ('10855101'), ('10860100'), ('10861100'), ('10863100'),
        ('10864100'), ('10865102'), ('10867100'), ('10868001'), ('10873100')
    ) AS v(DepartmentEpicId)
),

-- Exact AOM ICD-10-CM inclusion:
-- H66.001-H66.019, H66.40-H66.43, and H66.90-H66.93.
-- Resolving DiagnosisKey first handles multiple terminology rows per code.
aom_diagnosis_keys AS (
    SELECT DISTINCT
        dt.DiagnosisKey
    FROM dbo.DiagnosisTerminologyDim AS dt
    WHERE dt.Type = 'ICD-10-CM'
      AND (
            dt.Value LIKE 'H66.00[1-9]'
         OR dt.Value LIKE 'H66.01[0-9]'
         OR dt.Value IN (
                'H66.40', 'H66.41', 'H66.42', 'H66.43',
                'H66.90', 'H66.91', 'H66.92', 'H66.93'
         )
      )
),

-- One row per patient and qualifying AOM encounter. Retaining EncounterKey and
-- EncounterDate supports recurrence, antibiotic linkage, and follow-up tasks.
aom_encounters AS (
    SELECT DISTINCT
        def.PatientDurableKey,
        def.EncounterKey,
        CAST(ef.Date AS DATE) AS EncounterDate,
        dd.DepartmentEpicId
    FROM dbo.DiagnosisEventFact AS def
    INNER JOIN aom_diagnosis_keys AS aom
        ON aom.DiagnosisKey = def.DiagnosisKey

    -- Require the diagnosis to be attached to a real encounter.
    INNER JOIN dbo.EncounterFact AS ef
        ON ef.EncounterKey = def.EncounterKey

    -- Apply the supplied clinic list through the encounter's historical
    -- DepartmentKey and stable DepartmentEpicId.
    INNER JOIN dbo.DepartmentDim AS dd
```

```
        ON dd.DepartmentKey = ef.DepartmentKey
    INNER JOIN smch_pediatric_departments AS smch
        ON smch.DepartmentEpicId = dd.DepartmentEpicId

    -- PatientDim is history-tracked. Use exactly one current patient record.
    INNER JOIN dbo.PatientDim AS p
        ON p.DurableKey = def.PatientDurableKey
       AND p.IsCurrent = 1

    -- Diagnosis source: encounter diagnosis only. Explicitly exclude billing
    -- diagnoses, medical history, and problem-list diagnoses.
    WHERE def.Type = 'Encounter Diagnosis'

      -- Completed in-person outpatient encounter.
      -- The Caboodle face-to-face flag alone included telehealth/video visits,
      -- so also exclude video flags and virtual/phone VisitType labels.
      AND ef.DerivedEncounterStatus = 'Complete'
      AND ef.IsOutpatientFaceToFaceVisit = 1
      AND COALESCE(ef.IsVideoVisit, 0) = 0
      AND UPPER(COALESCE(ef.VisitType, '')) NOT LIKE '%TELEHEALTH%'
      AND UPPER(COALESCE(ef.VisitType, '')) NOT LIKE '%VIRTUAL%'
      AND UPPER(COALESCE(ef.VisitType, '')) NOT LIKE '%PHONE%'

      -- Inclusive calendar-date window that remains correct when ef.Date
      -- contains a time component.
      AND ef.Date >= @AomStartDate
      AND ef.Date < DATEADD(DAY, 1, @AomEndDate)

      -- Younger than 26 on the encounter date. This direct birthday comparison
      -- excludes future birth dates and excludes patients on/after age 26.
      -- Approved leap-day convention: SQL Server treats February 28 as the
      -- birthday in a non-leap year; no validated patients were affected.
      AND p.BirthDate IS NOT NULL
      AND p.BirthDate <= CAST(ef.Date AS DATE)
      AND CAST(ef.Date AS DATE) < DATEADD(YEAR, 26, p.BirthDate)

      -- Valid analytic patient only. These safeguards changed neither the
      -- validated index cohort nor the look-back population.
      AND p.IsValid = 1
      AND p.Test = 0
      AND p.DurableKey > 0
)

/* END SHARED DEFINITION: definitions/AOM-ENCOUNTER.sql */

/* BEGIN SHARED DEFINITION: definitions/AOM-ANTIBIOTIC.sql (AOM-ABX v0.3)
   CTEs: systemic_antibiotic_order_base, systemic_antibiotic_orders,
         aom_antibiotic_order_candidates, aom_antibiotic_orders,
         first_aom_antibiotic_per_patient, antibiotic_treated_aom_cohort */

, systemic_antibiotic_order_base AS (
    SELECT
        mo.MedicationOrderKey,
        mo.PatientDurableKey,
```

```
        mo.EncounterKey AS OrderEncounterKey,
        mo.OrderedInstant,
        LOWER(md.SimpleGenericName) AS NormalizedIngredientName
    FROM dbo.MedicationOrderFact AS mo
    INNER JOIN dbo.MedicationDim AS md
        ON md.MedicationKey = mo.MedicationKey
    INNER JOIN dbo.DepartmentDim AS department
        ON department.DepartmentKey = mo.DepartmentKey
    INNER JOIN smch_pediatric_departments AS clinic
        ON clinic.DepartmentEpicId = department.DepartmentEpicId
    WHERE mo.Type = 'Medications'
      AND mo.IsPending = 0
      AND mo._IsDeleted = 0
      AND mo.OrderedInstant >= @StudyStartDate

      -- Keep return-treatment orders needed by Task 10. AOM linkage below
      -- remains restricted to Day 0 through Day 3.
      AND mo.OrderedInstant < DATEADD(DAY, 15, @StudyEndDate)
      AND UPPER(md.TherapeuticClass) = 'ANTIBIOTICS'
      AND UPPER(md.Route) IN ('PO', 'INJ', 'IV', 'IM', 'SUBQ', 'SC')
),
systemic_antibiotic_orders AS (
    SELECT
        base.MedicationOrderKey,
        base.PatientDurableKey,
        base.OrderEncounterKey,
        base.OrderedInstant,
        CASE
            WHEN base.NormalizedIngredientName LIKE '%amoxicillin%clav%'
                THEN 'amoxicillin-clavulanate'
            WHEN base.NormalizedIngredientName LIKE 'amoxicillin%'
                THEN 'amoxicillin'
            WHEN base.NormalizedIngredientName LIKE 'cefdinir%' THEN 'cefdinir'
            WHEN base.NormalizedIngredientName LIKE 'clindamycin%' THEN 'clindamycin'
            WHEN base.NormalizedIngredientName LIKE 'ceftriaxone%' THEN 'ceftriaxone'
            ELSE base.NormalizedIngredientName
        END AS AntibioticAgent
    FROM systemic_antibiotic_order_base AS base
),
aom_antibiotic_order_candidates AS (
    SELECT
        antibiotic.MedicationOrderKey,
        antibiotic.PatientDurableKey,
        aom.EncounterKey AS AomEncounterKey,
        aom.EncounterDate AS AomEncounterDate,
        antibiotic.OrderEncounterKey,
        antibiotic.OrderedInstant,
        antibiotic.AntibioticAgent,
        ROW_NUMBER() OVER (
            PARTITION BY
                antibiotic.PatientDurableKey,
                antibiotic.MedicationOrderKey
            ORDER BY aom.EncounterDate DESC, aom.EncounterKey DESC
        ) AS AomAttributionRank
```

```
    FROM systemic_antibiotic_orders AS antibiotic
    INNER JOIN aom_encounters AS aom
        ON aom.PatientDurableKey = antibiotic.PatientDurableKey
       AND antibiotic.OrderedInstant >= CAST(aom.EncounterDate AS DATETIME)
       AND antibiotic.OrderedInstant
            < DATEADD(DAY, 4, CAST(aom.EncounterDate AS DATETIME))
),
aom_antibiotic_orders AS (
    SELECT
        candidate.MedicationOrderKey,
        candidate.PatientDurableKey,
        candidate.AomEncounterKey,
        candidate.AomEncounterDate,
        candidate.OrderEncounterKey,
        candidate.OrderedInstant,
        candidate.AntibioticAgent,
        CASE
            WHEN candidate.AntibioticAgent IN (
                'amoxicillin',
                'amoxicillin-clavulanate',
                'cefdinir',
                'clindamycin',
                'ceftriaxone'
            ) THEN candidate.AntibioticAgent
            ELSE 'other'
        END AS NamedAntibioticCategory,
        CASE
            WHEN candidate.AntibioticAgent = 'amoxicillin'
                THEN 'narrow-spectrum'
            WHEN candidate.AntibioticAgent IN (
                'amoxicillin-clavulanate',
                'cefdinir',
                'clindamycin',
                'ceftriaxone'
            ) THEN 'broad-spectrum'
            ELSE 'other'
        END AS SpectrumCategory
    FROM aom_antibiotic_order_candidates AS candidate
    WHERE candidate.AomAttributionRank = 1
),
first_aom_antibiotic_per_patient AS (
    SELECT
        linked.*,
        ROW_NUMBER() OVER (
            PARTITION BY linked.PatientDurableKey
            ORDER BY linked.OrderedInstant, linked.MedicationOrderKey
        ) AS FirstAntibioticRank
    FROM aom_antibiotic_orders AS linked
),
antibiotic_treated_aom_cohort AS (
    SELECT
        first_order.PatientDurableKey,
        first_order.AomEncounterKey AS IndexAomEncounterKey,
        first_order.AomEncounterDate AS IndexAomEncounterDate,
```

```
        first_order.MedicationOrderKey AS FirstMedicationOrderKey,
        first_order.OrderEncounterKey AS FirstAntibioticOrderEncounterKey,
        first_order.OrderedInstant AS FirstAntibioticOrderedInstant,
        first_order.AntibioticAgent AS FirstAntibioticAgent,
        first_order.NamedAntibioticCategory AS FirstNamedAntibioticCategory,
        first_order.SpectrumCategory AS FirstSpectrumCategory
    FROM first_aom_antibiotic_per_patient AS first_order
    WHERE first_order.FirstAntibioticRank = 1
)

/* END SHARED DEFINITION: definitions/AOM-ANTIBIOTIC.sql */

/* BEGIN TASK-SPECIFIC LOGIC: Task 4 */

SELECT
    (SELECT COUNT(*) FROM antibiotic_treated_aom_cohort)
        AS PatientsWithSystemicAntibioticWithin3Days,
    (SELECT COUNT(DISTINCT ae.PatientDurableKey) FROM aom_encounters AS ae)
        AS AomCohortPatients;

/* END TASK-SPECIFIC LOGIC: Task 4 */
```

### Task 5. Antibiotic Frequency

```
/*
EXECUTION RESULT
Run date: 2026-07-31 (America/Los_Angeles)
Command: uv run python query.py -f .\queries\05_frequency_of_5_named_antibiotics.sql

AntibioticCategory          PatientCount  Percent  Denominator
amoxicillin                         8595    80.91        10623
amoxicillin-clavulanate             2177    20.49        10623
cefdinir                            1384    13.03        10623
clindamycin                           13     0.12        10623
ceftriaxone                          187     1.76        10623
other                                573     5.39        10623
*/

/*
Task 5: Frequency of five named antibiotics

Shared definitions copied below:
- definitions/AOM-ENCOUNTER.sql
- definitions/AOM-ANTIBIOTIC.sql
- A patient is counted once per category but may occur in multiple categories.
*/

DECLARE @StudyStartDate DATE = '2023-01-01';
DECLARE @StudyEndDate   DATE = '2024-12-31';
DECLARE @AomStartDate   DATE = @StudyStartDate;
DECLARE @AomEndDate     DATE = @StudyEndDate;
```

```sql
/* BEGIN SHARED DEFINITION: definitions/AOM-ENCOUNTER.sql (AOM-ENC v1.0)
   CTEs: smch_pediatric_departments, aom_diagnosis_keys, aom_encounters */

-- Exact protocol-supplied SMCH primary care pediatric departments.
-- All 30 IDs mapped and had utilization during validation. Keep the explicitly
-- supplied ZZZ-named historical IDs; do not infer an additional active-status
-- restriction from the department name.
WITH smch_pediatric_departments AS (
    SELECT v.DepartmentEpicId
    FROM (VALUES
        ('10801100'), ('10802100'), ('10803100'), ('10804100'), ('10806100'),
        ('10807100'), ('10808100'), ('10809100'), ('10810100'), ('10814100'),
        ('10815100'), ('10816100'), ('10829100'), ('10830100'), ('10837100'),
        ('10838100'), ('10839100'), ('10842100'), ('10843100'), ('10847100'),
        ('10850105'), ('10855101'), ('10860100'), ('10861100'), ('10863100'),
        ('10864100'), ('10865102'), ('10867100'), ('10868001'), ('10873100')
    ) AS v(DepartmentEpicId)
),

-- Exact AOM ICD-10-CM inclusion:
-- H66.001-H66.019, H66.40-H66.43, and H66.90-H66.93.
-- Resolving DiagnosisKey first handles multiple terminology rows per code.
aom_diagnosis_keys AS (
    SELECT DISTINCT
        dt.DiagnosisKey
    FROM dbo.DiagnosisTerminologyDim AS dt
    WHERE dt.Type = 'ICD-10-CM'
      AND (
            dt.Value LIKE 'H66.00[1-9]'
         OR dt.Value LIKE 'H66.01[0-9]'
         OR dt.Value IN (
                'H66.40', 'H66.41', 'H66.42', 'H66.43',
                'H66.90', 'H66.91', 'H66.92', 'H66.93'
         )
      )
),

-- One row per patient and qualifying AOM encounter. Retaining EncounterKey and
-- EncounterDate supports recurrence, antibiotic linkage, and follow-up tasks.
aom_encounters AS (
    SELECT DISTINCT
        def.PatientDurableKey,
        def.EncounterKey,
        CAST(ef.Date AS DATE) AS EncounterDate,
        dd.DepartmentEpicId
    FROM dbo.DiagnosisEventFact AS def
    INNER JOIN aom_diagnosis_keys AS aom
        ON aom.DiagnosisKey = def.DiagnosisKey

    -- Require the diagnosis to be attached to a real encounter.
    INNER JOIN dbo.EncounterFact AS ef
        ON ef.EncounterKey = def.EncounterKey

    -- Apply the supplied clinic list through the encounter's historical
```

```sql
    -- DepartmentKey and stable DepartmentEpicId.
    INNER JOIN dbo.DepartmentDim AS dd
        ON dd.DepartmentKey = ef.DepartmentKey
    INNER JOIN smch_pediatric_departments AS smch
        ON smch.DepartmentEpicId = dd.DepartmentEpicId

    -- PatientDim is history-tracked. Use exactly one current patient record.
    INNER JOIN dbo.PatientDim AS p
        ON p.DurableKey = def.PatientDurableKey
       AND p.IsCurrent = 1

    -- Diagnosis source: encounter diagnosis only. Explicitly exclude billing
    -- diagnoses, medical history, and problem-list diagnoses.
    WHERE def.Type = 'Encounter Diagnosis'

      -- Completed in-person outpatient encounter.
      -- The Caboodle face-to-face flag alone included telehealth/video visits,
      -- so also exclude video flags and virtual/phone VisitType labels.
      AND ef.DerivedEncounterStatus = 'Complete'
      AND ef.IsOutpatientFaceToFaceVisit = 1
      AND COALESCE(ef.IsVideoVisit, 0) = 0
      AND UPPER(COALESCE(ef.VisitType, '')) NOT LIKE '%TELEHEALTH%'
      AND UPPER(COALESCE(ef.VisitType, '')) NOT LIKE '%VIRTUAL%'
      AND UPPER(COALESCE(ef.VisitType, '')) NOT LIKE '%PHONE%'

      -- Inclusive calendar-date window that remains correct when ef.Date
      -- contains a time component.
      AND ef.Date >= @AomStartDate
      AND ef.Date < DATEADD(DAY, 1, @AomEndDate)

      -- Younger than 26 on the encounter date. This direct birthday comparison
      -- excludes future birth dates and excludes patients on/after age 26.
      -- Approved leap-day convention: SQL Server treats February 28 as the
      -- birthday in a non-leap year; no validated patients were affected.
      AND p.BirthDate IS NOT NULL
      AND p.BirthDate <= CAST(ef.Date AS DATE)
      AND CAST(ef.Date AS DATE) < DATEADD(YEAR, 26, p.BirthDate)

      -- Valid analytic patient only. These safeguards changed neither the
      -- validated index cohort nor the look-back population.
      AND p.IsValid = 1
      AND p.Test = 0
      AND p.DurableKey > 0
)

/* END SHARED DEFINITION: definitions/AOM-ENCOUNTER.sql */

/* BEGIN SHARED DEFINITION: definitions/AOM-ANTIBIOTIC.sql (AOM-ABX v0.3)
   CTEs: systemic_antibiotic_order_base, systemic_antibiotic_orders,
         aom_antibiotic_order_candidates, aom_antibiotic_orders,
         first_aom_antibiotic_per_patient, antibiotic_treated_aom_cohort */

, systemic_antibiotic_order_base AS (
    SELECT
```

```
        mo.MedicationOrderKey,
        mo.PatientDurableKey,
        mo.EncounterKey AS OrderEncounterKey,
        mo.OrderedInstant,
        LOWER(md.SimpleGenericName) AS NormalizedIngredientName
    FROM dbo.MedicationOrderFact AS mo
    INNER JOIN dbo.MedicationDim AS md
        ON md.MedicationKey = mo.MedicationKey
    INNER JOIN dbo.DepartmentDim AS department
        ON department.DepartmentKey = mo.DepartmentKey
    INNER JOIN smch_pediatric_departments AS clinic
        ON clinic.DepartmentEpicId = department.DepartmentEpicId
    WHERE mo.Type = 'Medications'
      AND mo.IsPending = 0
      AND mo._IsDeleted = 0
      AND mo.OrderedInstant >= @StudyStartDate

      -- Keep return-treatment orders needed by Task 10. AOM linkage below
      -- remains restricted to Day 0 through Day 3.
      AND mo.OrderedInstant < DATEADD(DAY, 15, @StudyEndDate)
      AND UPPER(md.TherapeuticClass) = 'ANTIBIOTICS'
      AND UPPER(md.Route) IN ('PO', 'INJ', 'IV', 'IM', 'SUBQ', 'SC')
),
systemic_antibiotic_orders AS (
    SELECT
        base.MedicationOrderKey,
        base.PatientDurableKey,
        base.OrderEncounterKey,
        base.OrderedInstant,
        CASE
            WHEN base.NormalizedIngredientName LIKE '%amoxicillin%clav%'
                THEN 'amoxicillin-clavulanate'
            WHEN base.NormalizedIngredientName LIKE 'amoxicillin%'
                THEN 'amoxicillin'
            WHEN base.NormalizedIngredientName LIKE 'cefdinir%' THEN 'cefdinir'
            WHEN base.NormalizedIngredientName LIKE 'clindamycin%' THEN 'clindamycin'
            WHEN base.NormalizedIngredientName LIKE 'ceftriaxone%' THEN 'ceftriaxone'
            ELSE base.NormalizedIngredientName
        END AS AntibioticAgent
    FROM systemic_antibiotic_order_base AS base
),
aom_antibiotic_order_candidates AS (
    SELECT
        antibiotic.MedicationOrderKey,
        antibiotic.PatientDurableKey,
        aom.EncounterKey AS AomEncounterKey,
        aom.EncounterDate AS AomEncounterDate,
        antibiotic.OrderEncounterKey,
        antibiotic.OrderedInstant,
        antibiotic.AntibioticAgent,
        ROW_NUMBER() OVER (
            PARTITION BY
                antibiotic.PatientDurableKey,
                antibiotic.MedicationOrderKey
```

```
            ORDER BY aom.EncounterDate DESC, aom.EncounterKey DESC
        ) AS AomAttributionRank
    FROM systemic_antibiotic_orders AS antibiotic
    INNER JOIN aom_encounters AS aom
        ON aom.PatientDurableKey = antibiotic.PatientDurableKey
       AND antibiotic.OrderedInstant >= CAST(aom.EncounterDate AS DATETIME)
       AND antibiotic.OrderedInstant
            < DATEADD(DAY, 4, CAST(aom.EncounterDate AS DATETIME))
),
aom_antibiotic_orders AS (
    SELECT
        candidate.MedicationOrderKey,
        candidate.PatientDurableKey,
        candidate.AomEncounterKey,
        candidate.AomEncounterDate,
        candidate.OrderEncounterKey,
        candidate.OrderedInstant,
        candidate.AntibioticAgent,
        CASE
            WHEN candidate.AntibioticAgent IN (
                'amoxicillin',
                'amoxicillin-clavulanate',
                'cefdinir',
                'clindamycin',
                'ceftriaxone'
            ) THEN candidate.AntibioticAgent
            ELSE 'other'
        END AS NamedAntibioticCategory,
        CASE
            WHEN candidate.AntibioticAgent = 'amoxicillin'
                THEN 'narrow-spectrum'
            WHEN candidate.AntibioticAgent IN (
                'amoxicillin-clavulanate',
                'cefdinir',
                'clindamycin',
                'ceftriaxone'
            ) THEN 'broad-spectrum'
            ELSE 'other'
        END AS SpectrumCategory
    FROM aom_antibiotic_order_candidates AS candidate
    WHERE candidate.AomAttributionRank = 1
),
first_aom_antibiotic_per_patient AS (
    SELECT
        linked.*,
        ROW_NUMBER() OVER (
            PARTITION BY linked.PatientDurableKey
            ORDER BY linked.OrderedInstant, linked.MedicationOrderKey
        ) AS FirstAntibioticRank
    FROM aom_antibiotic_orders AS linked
),
antibiotic_treated_aom_cohort AS (
    SELECT
        first_order.PatientDurableKey,
```

```sql
        first_order.AomEncounterKey AS IndexAomEncounterKey,
        first_order.AomEncounterDate AS IndexAomEncounterDate,
        first_order.MedicationOrderKey AS FirstMedicationOrderKey,
        first_order.OrderEncounterKey AS FirstAntibioticOrderEncounterKey,
        first_order.OrderedInstant AS FirstAntibioticOrderedInstant,
        first_order.AntibioticAgent AS FirstAntibioticAgent,
        first_order.NamedAntibioticCategory AS FirstNamedAntibioticCategory,
        first_order.SpectrumCategory AS FirstSpectrumCategory
    FROM first_aom_antibiotic_per_patient AS first_order
    WHERE first_order.FirstAntibioticRank = 1
)

/* END SHARED DEFINITION: definitions/AOM-ANTIBIOTIC.sql */

/* BEGIN TASK-SPECIFIC LOGIC: Task 5 */

, antibiotic_categories AS (
    SELECT v.AntibioticCategory, v.SortOrder
    FROM (VALUES
        ('amoxicillin', 1),
        ('amoxicillin-clavulanate', 2),
        ('cefdinir', 3),
        ('clindamycin', 4),
        ('ceftriaxone', 5),
        ('other', 6)
    ) AS v(AntibioticCategory, SortOrder)
),
patient_antibiotic_categories AS (
    SELECT DISTINCT
        linked.PatientDurableKey,
        linked.NamedAntibioticCategory AS AntibioticCategory
    FROM aom_antibiotic_orders AS linked
),
category_counts AS (
    SELECT
        category.AntibioticCategory,
        category.SortOrder,
        COUNT(patient_category.PatientDurableKey) AS PatientCount
    FROM antibiotic_categories AS category
    LEFT JOIN patient_antibiotic_categories AS patient_category
        ON patient_category.AntibioticCategory = category.AntibioticCategory
    GROUP BY
        category.AntibioticCategory,
        category.SortOrder
),
cohort_size AS (
    SELECT COUNT(*) AS AntibioticTreatedCohortPatients
    FROM antibiotic_treated_aom_cohort
)
SELECT
    counts.AntibioticCategory,
    counts.PatientCount,
    CAST(
        100.0 * counts.PatientCount
```

```
        / NULLIF(cohort.AntibioticTreatedCohortPatients, 0)
        AS DECIMAL(6, 2)
    ) AS PercentOfAntibioticTreatedCohort,
    cohort.AntibioticTreatedCohortPatients
FROM category_counts AS counts
CROSS JOIN cohort_size AS cohort
ORDER BY counts.SortOrder;

/* END TASK-SPECIFIC LOGIC: Task 5 */
```

### Task 6. Antibiotic Spectrum Classification

```
/*
EXECUTION RESULT
Run date: 2026-07-31 (America/Los_Angeles)
Command: uv run python query.py -f .\queries\06_first_antibiotic_spectrum_proportion.sql

SpectrumCategory  PatientCount  Percent  Denominator
narrow-spectrum           8225    77.43        10623
broad-spectrum            2068    19.47        10623
other                      330     3.11        10623
*/

/*
Task 6: Spectrum of first antibiotic

Shared definitions copied below:
- definitions/AOM-ENCOUNTER.sql
- definitions/AOM-ANTIBIOTIC.sql
- AOM-ANTIBIOTIC supplies one deterministic first antibiotic per patient.
*/

DECLARE @StudyStartDate DATE = '2023-01-01';
DECLARE @StudyEndDate   DATE = '2024-12-31';
DECLARE @AomStartDate   DATE = @StudyStartDate;
DECLARE @AomEndDate     DATE = @StudyEndDate;

/* BEGIN SHARED DEFINITION: definitions/AOM-ENCOUNTER.sql (AOM-ENC v1.0)
   CTEs: smch_pediatric_departments, aom_diagnosis_keys, aom_encounters */

-- Exact protocol-supplied SMCH primary care pediatric departments.
-- All 30 IDs mapped and had utilization during validation. Keep the explicitly
-- supplied ZZZ-named historical IDs; do not infer an additional active-status
-- restriction from the department name.
WITH smch_pediatric_departments AS (
    SELECT v.DepartmentEpicId
    FROM (VALUES
        ('10801100'), ('10802100'), ('10803100'), ('10804100'), ('10806100'),
        ('10807100'), ('10808100'), ('10809100'), ('10810100'), ('10814100'),
        ('10815100'), ('10816100'), ('10829100'), ('10830100'), ('10837100'),
        ('10838100'), ('10839100'), ('10842100'), ('10843100'), ('10847100'),
        ('10850105'), ('10855101'), ('10860100'), ('10861100'), ('10863100'),
```

```sql
        ('10864100'), ('10865102'), ('10867100'), ('10868001'), ('10873100')
    ) AS v(DepartmentEpicId)
),

-- Exact AOM ICD-10-CM inclusion:
-- H66.001-H66.019, H66.40-H66.43, and H66.90-H66.93.
-- Resolving DiagnosisKey first handles multiple terminology rows per code.
aom_diagnosis_keys AS (
    SELECT DISTINCT
        dt.DiagnosisKey
    FROM dbo.DiagnosisTerminologyDim AS dt
    WHERE dt.Type = 'ICD-10-CM'
      AND (
            dt.Value LIKE 'H66.00[1-9]'
         OR dt.Value LIKE 'H66.01[0-9]'
         OR dt.Value IN (
                'H66.40', 'H66.41', 'H66.42', 'H66.43',
                'H66.90', 'H66.91', 'H66.92', 'H66.93'
         )
      )
),

-- One row per patient and qualifying AOM encounter. Retaining EncounterKey and
-- EncounterDate supports recurrence, antibiotic linkage, and follow-up tasks.
aom_encounters AS (
    SELECT DISTINCT
        def.PatientDurableKey,
        def.EncounterKey,
        CAST(ef.Date AS DATE) AS EncounterDate,
        dd.DepartmentEpicId
    FROM dbo.DiagnosisEventFact AS def
    INNER JOIN aom_diagnosis_keys AS aom
        ON aom.DiagnosisKey = def.DiagnosisKey

    -- Require the diagnosis to be attached to a real encounter.
    INNER JOIN dbo.EncounterFact AS ef
        ON ef.EncounterKey = def.EncounterKey

    -- Apply the supplied clinic list through the encounter's historical
    -- DepartmentKey and stable DepartmentEpicId.
    INNER JOIN dbo.DepartmentDim AS dd
        ON dd.DepartmentKey = ef.DepartmentKey
    INNER JOIN smch_pediatric_departments AS smch
        ON smch.DepartmentEpicId = dd.DepartmentEpicId

    -- PatientDim is history-tracked. Use exactly one current patient record.
    INNER JOIN dbo.PatientDim AS p
        ON p.DurableKey = def.PatientDurableKey
       AND p.IsCurrent = 1

    -- Diagnosis source: encounter diagnosis only. Explicitly exclude billing
    -- diagnoses, medical history, and problem-list diagnoses.
    WHERE def.Type = 'Encounter Diagnosis'
```

```sql
      -- Completed in-person outpatient encounter.
      -- The Caboodle face-to-face flag alone included telehealth/video visits,
      -- so also exclude video flags and virtual/phone VisitType labels.
      AND ef.DerivedEncounterStatus = 'Complete'
      AND ef.IsOutpatientFaceToFaceVisit = 1
      AND COALESCE(ef.IsVideoVisit, 0) = 0
      AND UPPER(COALESCE(ef.VisitType, '')) NOT LIKE '%TELEHEALTH%'
      AND UPPER(COALESCE(ef.VisitType, '')) NOT LIKE '%VIRTUAL%'
      AND UPPER(COALESCE(ef.VisitType, '')) NOT LIKE '%PHONE%'

      -- Inclusive calendar-date window that remains correct when ef.Date
      -- contains a time component.
      AND ef.Date >= @AomStartDate
      AND ef.Date < DATEADD(DAY, 1, @AomEndDate)

      -- Younger than 26 on the encounter date. This direct birthday comparison
      -- excludes future birth dates and excludes patients on/after age 26.
      -- Approved leap-day convention: SQL Server treats February 28 as the
      -- birthday in a non-leap year; no validated patients were affected.
      AND p.BirthDate IS NOT NULL
      AND p.BirthDate <= CAST(ef.Date AS DATE)
      AND CAST(ef.Date AS DATE) < DATEADD(YEAR, 26, p.BirthDate)

      -- Valid analytic patient only. These safeguards changed neither the
      -- validated index cohort nor the look-back population.
      AND p.IsValid = 1
      AND p.Test = 0
      AND p.DurableKey > 0
)

/* END SHARED DEFINITION: definitions/AOM-ENCOUNTER.sql */

/* BEGIN SHARED DEFINITION: definitions/AOM-ANTIBIOTIC.sql (AOM-ABX v0.3)
   CTEs: systemic_antibiotic_order_base, systemic_antibiotic_orders,
         aom_antibiotic_order_candidates, aom_antibiotic_orders,
         first_aom_antibiotic_per_patient, antibiotic_treated_aom_cohort */

, systemic_antibiotic_order_base AS (
    SELECT
        mo.MedicationOrderKey,
        mo.PatientDurableKey,
        mo.EncounterKey AS OrderEncounterKey,
        mo.OrderedInstant,
        LOWER(md.SimpleGenericName) AS NormalizedIngredientName
    FROM dbo.MedicationOrderFact AS mo
    INNER JOIN dbo.MedicationDim AS md
        ON md.MedicationKey = mo.MedicationKey
    INNER JOIN dbo.DepartmentDim AS department
        ON department.DepartmentKey = mo.DepartmentKey
    INNER JOIN smch_pediatric_departments AS clinic
        ON clinic.DepartmentEpicId = department.DepartmentEpicId
    WHERE mo.Type = 'Medications'
      AND mo.IsPending = 0
      AND mo._IsDeleted = 0
```

```sql
      AND mo.OrderedInstant >= @StudyStartDate

      -- Keep return-treatment orders needed by Task 10. AOM linkage below
      -- remains restricted to Day 0 through Day 3.
      AND mo.OrderedInstant < DATEADD(DAY, 15, @StudyEndDate)
      AND UPPER(md.TherapeuticClass) = 'ANTIBIOTICS'
      AND UPPER(md.Route) IN ('PO', 'INJ', 'IV', 'IM', 'SUBQ', 'SC')
),
systemic_antibiotic_orders AS (
    SELECT
        base.MedicationOrderKey,
        base.PatientDurableKey,
        base.OrderEncounterKey,
        base.OrderedInstant,
        CASE
            WHEN base.NormalizedIngredientName LIKE '%amoxicillin%clav%'
                THEN 'amoxicillin-clavulanate'
            WHEN base.NormalizedIngredientName LIKE 'amoxicillin%'
                THEN 'amoxicillin'
            WHEN base.NormalizedIngredientName LIKE 'cefdinir%' THEN 'cefdinir'
            WHEN base.NormalizedIngredientName LIKE 'clindamycin%' THEN 'clindamycin'
            WHEN base.NormalizedIngredientName LIKE 'ceftriaxone%' THEN 'ceftriaxone'
            ELSE base.NormalizedIngredientName
        END AS AntibioticAgent
    FROM systemic_antibiotic_order_base AS base
),
aom_antibiotic_order_candidates AS (
    SELECT
        antibiotic.MedicationOrderKey,
        antibiotic.PatientDurableKey,
        aom.EncounterKey AS AomEncounterKey,
        aom.EncounterDate AS AomEncounterDate,
        antibiotic.OrderEncounterKey,
        antibiotic.OrderedInstant,
        antibiotic.AntibioticAgent,
        ROW_NUMBER() OVER (
            PARTITION BY
                antibiotic.PatientDurableKey,
                antibiotic.MedicationOrderKey
            ORDER BY aom.EncounterDate DESC, aom.EncounterKey DESC
        ) AS AomAttributionRank
    FROM systemic_antibiotic_orders AS antibiotic
    INNER JOIN aom_encounters AS aom
        ON aom.PatientDurableKey = antibiotic.PatientDurableKey
       AND antibiotic.OrderedInstant >= CAST(aom.EncounterDate AS DATETIME)
       AND antibiotic.OrderedInstant
            < DATEADD(DAY, 4, CAST(aom.EncounterDate AS DATETIME))
),
aom_antibiotic_orders AS (
    SELECT
        candidate.MedicationOrderKey,
        candidate.PatientDurableKey,
        candidate.AomEncounterKey,
        candidate.AomEncounterDate,
```

```
        candidate.OrderEncounterKey,
        candidate.OrderedInstant,
        candidate.AntibioticAgent,
        CASE
            WHEN candidate.AntibioticAgent IN (
                'amoxicillin',
                'amoxicillin-clavulanate',
                'cefdinir',
                'clindamycin',
                'ceftriaxone'
            ) THEN candidate.AntibioticAgent
            ELSE 'other'
        END AS NamedAntibioticCategory,
        CASE
            WHEN candidate.AntibioticAgent = 'amoxicillin'
                THEN 'narrow-spectrum'
            WHEN candidate.AntibioticAgent IN (
                'amoxicillin-clavulanate',
                'cefdinir',
                'clindamycin',
                'ceftriaxone'
            ) THEN 'broad-spectrum'
            ELSE 'other'
        END AS SpectrumCategory
    FROM aom_antibiotic_order_candidates AS candidate
    WHERE candidate.AomAttributionRank = 1
),
first_aom_antibiotic_per_patient AS (
    SELECT
        linked.*,
        ROW_NUMBER() OVER (
            PARTITION BY linked.PatientDurableKey
            ORDER BY linked.OrderedInstant, linked.MedicationOrderKey
        ) AS FirstAntibioticRank
    FROM aom_antibiotic_orders AS linked
),
antibiotic_treated_aom_cohort AS (
    SELECT
        first_order.PatientDurableKey,
        first_order.AomEncounterKey AS IndexAomEncounterKey,
        first_order.AomEncounterDate AS IndexAomEncounterDate,
        first_order.MedicationOrderKey AS FirstMedicationOrderKey,
        first_order.OrderEncounterKey AS FirstAntibioticOrderEncounterKey,
        first_order.OrderedInstant AS FirstAntibioticOrderedInstant,
        first_order.AntibioticAgent AS FirstAntibioticAgent,
        first_order.NamedAntibioticCategory AS FirstNamedAntibioticCategory,
        first_order.SpectrumCategory AS FirstSpectrumCategory
    FROM first_aom_antibiotic_per_patient AS first_order
    WHERE first_order.FirstAntibioticRank = 1
)

/* END SHARED DEFINITION: definitions/AOM-ANTIBIOTIC.sql */

/* BEGIN TASK-SPECIFIC LOGIC: Task 6 */
```

```
, spectrum_categories AS (
    SELECT v.SpectrumCategory, v.SortOrder
    FROM (VALUES
        ('narrow-spectrum', 1),
        ('broad-spectrum', 2),
        ('other', 3)
    ) AS v(SpectrumCategory, SortOrder)
),
category_counts AS (
    SELECT
        category.SpectrumCategory,
        category.SortOrder,
        COUNT(cohort.PatientDurableKey) AS PatientCount
    FROM spectrum_categories AS category
    LEFT JOIN antibiotic_treated_aom_cohort AS cohort
        ON cohort.FirstSpectrumCategory = category.SpectrumCategory
    GROUP BY
        category.SpectrumCategory,
        category.SortOrder
),
cohort_size AS (
    SELECT COUNT(*) AS AntibioticTreatedCohortPatients
    FROM antibiotic_treated_aom_cohort
)
SELECT
    counts.SpectrumCategory,
    counts.PatientCount,
    CAST(
        100.0 * counts.PatientCount
        / NULLIF(cohort.AntibioticTreatedCohortPatients, 0)
        AS DECIMAL(6, 2)
    ) AS PercentOfAntibioticTreatedCohort,
    cohort.AntibioticTreatedCohortPatients
FROM category_counts AS counts
CROSS JOIN cohort_size AS cohort
ORDER BY counts.SortOrder;

/* END TASK-SPECIFIC LOGIC: Task 6 */
```

### Task 7. Tubes (Recurrent)

```
/*
EXECUTION RESULT
Run date: 2026-07-31 (America/Los_Angeles)
Command: uv run python query.py -f .\queries\07_recurrent_aom_with_tympanostomy_tubes.sql

RecurrentAomPatientsWithTubes: 55
RecurrentAomCohortPatients:    495
*/

/*
```

```
Task 7: Tympanostomy tubes among recurrent patients

Shared definitions copied below:
- AOM-ENCOUNTER uses the 2022 look-back start for recurrence.
- AOM-TYMPANOSTOMY still derives first AOM from the 2023-2024 index window.
*/

DECLARE @LookbackStartDate DATE = '2022-01-01';
DECLARE @StudyStartDate    DATE = '2023-01-01';
DECLARE @StudyEndDate      DATE = '2024-12-31';
DECLARE @FollowupEndDate   DATE = '2025-12-31';
DECLARE @AomStartDate      DATE = @LookbackStartDate;
DECLARE @AomEndDate        DATE = @StudyEndDate;

/* BEGIN SHARED DEFINITION: definitions/AOM-ENCOUNTER.sql (AOM-ENC v1.0)
   CTEs: smch_pediatric_departments, aom_diagnosis_keys, aom_encounters */

-- Exact protocol-supplied SMCH primary care pediatric departments.
-- All 30 IDs mapped and had utilization during validation. Keep the explicitly
-- supplied ZZZ-named historical IDs; do not infer an additional active-status
-- restriction from the department name.
WITH smch_pediatric_departments AS (
    SELECT v.DepartmentEpicId
    FROM (VALUES
        ('10801100'), ('10802100'), ('10803100'), ('10804100'), ('10806100'),
        ('10807100'), ('10808100'), ('10809100'), ('10810100'), ('10814100'),
        ('10815100'), ('10816100'), ('10829100'), ('10830100'), ('10837100'),
        ('10838100'), ('10839100'), ('10842100'), ('10843100'), ('10847100'),
        ('10850105'), ('10855101'), ('10860100'), ('10861100'), ('10863100'),
        ('10864100'), ('10865102'), ('10867100'), ('10868001'), ('10873100')
    ) AS v(DepartmentEpicId)
),

-- Exact AOM ICD-10-CM inclusion:
-- H66.001-H66.019, H66.40-H66.43, and H66.90-H66.93.
-- Resolving DiagnosisKey first handles multiple terminology rows per code.
aom_diagnosis_keys AS (
    SELECT DISTINCT
        dt.DiagnosisKey
    FROM dbo.DiagnosisTerminologyDim AS dt
    WHERE dt.Type = 'ICD-10-CM'
      AND (
            dt.Value LIKE 'H66.00[1-9]'
         OR dt.Value LIKE 'H66.01[0-9]'
         OR dt.Value IN (
                'H66.40', 'H66.41', 'H66.42', 'H66.43',
                'H66.90', 'H66.91', 'H66.92', 'H66.93'
         )
      )
),

-- One row per patient and qualifying AOM encounter. Retaining EncounterKey and
-- EncounterDate supports recurrence, antibiotic linkage, and follow-up tasks.
aom_encounters AS (
```

```sql
SELECT DISTINCT
    def.PatientDurableKey,
    def.EncounterKey,
    CAST(ef.Date AS DATE) AS EncounterDate,
    dd.DepartmentEpicId
FROM dbo.DiagnosisEventFact AS def
INNER JOIN aom_diagnosis_keys AS aom
    ON aom.DiagnosisKey = def.DiagnosisKey

-- Require the diagnosis to be attached to a real encounter.
INNER JOIN dbo.EncounterFact AS ef
    ON ef.EncounterKey = def.EncounterKey

-- Apply the supplied clinic list through the encounter's historical
-- DepartmentKey and stable DepartmentEpicId.
INNER JOIN dbo.DepartmentDim AS dd
    ON dd.DepartmentKey = ef.DepartmentKey
INNER JOIN smch_pediatric_departments AS smch
    ON smch.DepartmentEpicId = dd.DepartmentEpicId

-- PatientDim is history-tracked. Use exactly one current patient record.
INNER JOIN dbo.PatientDim AS p
    ON p.DurableKey = def.PatientDurableKey
   AND p.IsCurrent = 1

-- Diagnosis source: encounter diagnosis only. Explicitly exclude billing
-- diagnoses, medical history, and problem-list diagnoses.
WHERE def.Type = 'Encounter Diagnosis'

  -- Completed in-person outpatient encounter.
  -- The Caboodle face-to-face flag alone included telehealth/video visits,
  -- so also exclude video flags and virtual/phone VisitType labels.
  AND ef.DerivedEncounterStatus = 'Complete'
  AND ef.IsOutpatientFaceToFaceVisit = 1
  AND COALESCE(ef.IsVideoVisit, 0) = 0
  AND UPPER(COALESCE(ef.VisitType, '')) NOT LIKE '%TELEHEALTH%'
  AND UPPER(COALESCE(ef.VisitType, '')) NOT LIKE '%VIRTUAL%'
  AND UPPER(COALESCE(ef.VisitType, '')) NOT LIKE '%PHONE%'

  -- Inclusive calendar-date window that remains correct when ef.Date
  -- contains a time component.
  AND ef.Date >= @AomStartDate
  AND ef.Date < DATEADD(DAY, 1, @AomEndDate)

  -- Younger than 26 on the encounter date. This direct birthday comparison
  -- excludes future birth dates and excludes patients on/after age 26.
  -- Approved leap-day convention: SQL Server treats February 28 as the
  -- birthday in a non-leap year; no validated patients were affected.
  AND p.BirthDate IS NOT NULL
  AND p.BirthDate <= CAST(ef.Date AS DATE)
  AND CAST(ef.Date AS DATE) < DATEADD(YEAR, 26, p.BirthDate)

  -- Valid analytic patient only. These safeguards changed neither the
  -- validated index cohort nor the look-back population.
```

```
      AND p.IsValid = 1
      AND p.Test = 0
      AND p.DurableKey > 0
)

/* END SHARED DEFINITION: definitions/AOM-ENCOUNTER.sql */

/* BEGIN SHARED DEFINITION: definitions/AOM-RECURRENT.sql (AOM-REC v0.2)
   CTEs: aom_index_cohort, aom_episode_dates,
         candidate_recurrence_triplets, recurrent_aom_cohort */

, aom_index_cohort AS (
    SELECT DISTINCT
        ae.PatientDurableKey
    FROM aom_encounters AS ae
    WHERE ae.EncounterDate >= @StudyStartDate
      AND ae.EncounterDate < DATEADD(DAY, 1, @StudyEndDate)
),
aom_episode_dates AS (
    SELECT DISTINCT
        ae.PatientDurableKey,
        ae.EncounterDate AS EpisodeDate
    FROM aom_encounters AS ae
),
candidate_recurrence_triplets AS (
    SELECT
        first_episode.PatientDurableKey,
        first_episode.EpisodeDate AS Episode1Date,
        second_episode.EpisodeDate AS Episode2Date,
        third_episode.EpisodeDate AS Episode3Date
    FROM aom_episode_dates AS first_episode
    INNER JOIN aom_episode_dates AS second_episode
        ON second_episode.PatientDurableKey = first_episode.PatientDurableKey
       AND second_episode.EpisodeDate
            >= DATEADD(DAY, 30, first_episode.EpisodeDate)
    INNER JOIN aom_episode_dates AS third_episode
        ON third_episode.PatientDurableKey = second_episode.PatientDurableKey
       AND third_episode.EpisodeDate
            >= DATEADD(DAY, 30, second_episode.EpisodeDate)
       AND third_episode.EpisodeDate
            <= DATEADD(MONTH, 6, first_episode.EpisodeDate)
),
recurrent_aom_cohort AS (
    SELECT DISTINCT
        cohort.PatientDurableKey
    FROM aom_index_cohort AS cohort
    INNER JOIN candidate_recurrence_triplets AS triplet
        ON triplet.PatientDurableKey = cohort.PatientDurableKey
)

/* END SHARED DEFINITION: definitions/AOM-RECURRENT.sql */

/* BEGIN SHARED DEFINITION: definitions/AOM-TYMPANOSTOMY.sql (AOM-TUBE v0.3)
   CTEs: aom_index_first_dates, procedure_event_tube_dates,
```

```
         surgical_event_tube_dates, tympanostomy_dates,
         first_aom_tympanostomy */

, aom_index_first_dates AS (
    SELECT
        ae.PatientDurableKey,
        MIN(ae.EncounterDate) AS FirstAomDate
    FROM aom_encounters AS ae
    WHERE ae.EncounterDate >= @StudyStartDate
      AND ae.EncounterDate < DATEADD(DAY, 1, @StudyEndDate)
    GROUP BY ae.PatientDurableKey
),
procedure_event_tube_dates AS (
    SELECT DISTINCT
        pef.PatientDurableKey,
        CAST(pef.ProcedureStartInstant AS DATE) AS TubeDate
    FROM dbo.ProcedureEventFact AS pef
    WHERE pef._IsDeleted = 0
      AND COALESCE(pef.IsUnsuccessfulAttempt, 0) = 0
      AND pef.ProcedureStartInstant >= @StudyStartDate
      AND pef.ProcedureStartInstant < DATEADD(DAY, 1, @FollowupEndDate)
      AND pef.ProcedureCodeSet IN ('HcpcsCode', 'CPT(R)')
      AND pef.ProcedureCode IN ('69433', '69436', '0583T')
),
surgical_event_tube_dates AS (
    SELECT DISTINCT
        spef.PatientDurableKey,
        surgery_date.DateValue AS TubeDate
    FROM dbo.SurgicalProcedureEventFact AS spef
    INNER JOIN dbo.ProcedureDim AS procedure_code
        ON procedure_code.ProcedureKey = spef.ProcedureCodeKey
    INNER JOIN dbo.DateDim AS surgery_date
        ON surgery_date.DateKey = spef.SurgeryDateKey
    WHERE spef._IsDeleted = 0
      AND spef.Performed = 1
      AND surgery_date.DateValue >= @StudyStartDate
      AND surgery_date.DateValue < DATEADD(DAY, 1, @FollowupEndDate)
      AND (
            procedure_code.CptCode IN ('69433', '69436', '0583T')
         OR procedure_code.HcpcsCode IN ('69433', '69436', '0583T')
         OR procedure_code.Code IN ('69433', '69436', '0583T')
      )
),

-- UNION removes duplicate patient/date representations across both sources.
tympanostomy_dates AS (
    SELECT PatientDurableKey, TubeDate
    FROM procedure_event_tube_dates

    UNION

    SELECT PatientDurableKey, TubeDate
    FROM surgical_event_tube_dates
),
```

```sql
first_aom_tympanostomy AS (
    SELECT
        first_aom.PatientDurableKey,
        first_aom.FirstAomDate,
        MIN(tube.TubeDate) AS FirstTubeDate,
        DATEDIFF(DAY, first_aom.FirstAomDate, MIN(tube.TubeDate))
            AS DaysFirstAomToFirstTube
    FROM aom_index_first_dates AS first_aom
    INNER JOIN tympanostomy_dates AS tube
        ON tube.PatientDurableKey = first_aom.PatientDurableKey
       AND tube.TubeDate >= first_aom.FirstAomDate
    GROUP BY
        first_aom.PatientDurableKey,
        first_aom.FirstAomDate
)

/* END SHARED DEFINITION: definitions/AOM-TYMPANOSTOMY.sql */

/* BEGIN TASK-SPECIFIC LOGIC: Task 7 */

SELECT
    COUNT(tube.PatientDurableKey) AS RecurrentAomPatientsWithTubes,
    COUNT(*) AS RecurrentAomCohortPatients
FROM recurrent_aom_cohort AS recurrent
LEFT JOIN first_aom_tympanostomy AS tube
    ON tube.PatientDurableKey = recurrent.PatientDurableKey;

/* END TASK-SPECIFIC LOGIC: Task 7 */
```

### Task 8. Median Days to Tube

```sql
/*
EXECUTION RESULT
Run date: 2026-07-31 (America/Los_Angeles)
Command: uv run python query.py -f .\queries\08_median_days_first_aom_to_first_tube.sql

MedianDays:       208.00
Q1Days:           108.00
Q3Days:           362.00
IqrWidthDays:     254.00
PatientsIncluded: 237
*/

/*
Task 8: Time from first AOM to first tube

Shared definitions copied below:
- definitions/AOM-ENCOUNTER.sql
- definitions/AOM-TYMPANOSTOMY.sql
- AOM-TYMPANOSTOMY checks both ProcedureEventFact and
  SurgicalProcedureEventFact.
*/
```

```sql
DECLARE @StudyStartDate  DATE = '2023-01-01';
DECLARE @StudyEndDate    DATE = '2024-12-31';
DECLARE @FollowupEndDate DATE = '2025-12-31';
DECLARE @AomStartDate    DATE = @StudyStartDate;
DECLARE @AomEndDate      DATE = @StudyEndDate;

/* BEGIN SHARED DEFINITION: definitions/AOM-ENCOUNTER.sql (AOM-ENC v1.0)
   CTEs: smch_pediatric_departments, aom_diagnosis_keys, aom_encounters */

-- Exact protocol-supplied SMCH primary care pediatric departments.
-- All 30 IDs mapped and had utilization during validation. Keep the explicitly
-- supplied ZZZ-named historical IDs; do not infer an additional active-status
-- restriction from the department name.
WITH smch_pediatric_departments AS (
    SELECT v.DepartmentEpicId
    FROM (VALUES
        ('10801100'), ('10802100'), ('10803100'), ('10804100'), ('10806100'),
        ('10807100'), ('10808100'), ('10809100'), ('10810100'), ('10814100'),
        ('10815100'), ('10816100'), ('10829100'), ('10830100'), ('10837100'),
        ('10838100'), ('10839100'), ('10842100'), ('10843100'), ('10847100'),
        ('10850105'), ('10855101'), ('10860100'), ('10861100'), ('10863100'),
        ('10864100'), ('10865102'), ('10867100'), ('10868001'), ('10873100')
    ) AS v(DepartmentEpicId)
),

-- Exact AOM ICD-10-CM inclusion:
-- H66.001-H66.019, H66.40-H66.43, and H66.90-H66.93.
-- Resolving DiagnosisKey first handles multiple terminology rows per code.
aom_diagnosis_keys AS (
    SELECT DISTINCT
        dt.DiagnosisKey
    FROM dbo.DiagnosisTerminologyDim AS dt
    WHERE dt.Type = 'ICD-10-CM'
      AND (
            dt.Value LIKE 'H66.00[1-9]'
         OR dt.Value LIKE 'H66.01[0-9]'
         OR dt.Value IN (
                'H66.40', 'H66.41', 'H66.42', 'H66.43',
                'H66.90', 'H66.91', 'H66.92', 'H66.93'
         )
      )
),

-- One row per patient and qualifying AOM encounter. Retaining EncounterKey and
-- EncounterDate supports recurrence, antibiotic linkage, and follow-up tasks.
aom_encounters AS (
    SELECT DISTINCT
        def.PatientDurableKey,
        def.EncounterKey,
        CAST(ef.Date AS DATE) AS EncounterDate,
        dd.DepartmentEpicId
    FROM dbo.DiagnosisEventFact AS def
    INNER JOIN aom_diagnosis_keys AS aom
```

```sql
        ON aom.DiagnosisKey = def.DiagnosisKey

    -- Require the diagnosis to be attached to a real encounter.
    INNER JOIN dbo.EncounterFact AS ef
        ON ef.EncounterKey = def.EncounterKey

    -- Apply the supplied clinic list through the encounter's historical
    -- DepartmentKey and stable DepartmentEpicId.
    INNER JOIN dbo.DepartmentDim AS dd
        ON dd.DepartmentKey = ef.DepartmentKey
    INNER JOIN smch_pediatric_departments AS smch
        ON smch.DepartmentEpicId = dd.DepartmentEpicId

    -- PatientDim is history-tracked. Use exactly one current patient record.
    INNER JOIN dbo.PatientDim AS p
        ON p.DurableKey = def.PatientDurableKey
       AND p.IsCurrent = 1

    -- Diagnosis source: encounter diagnosis only. Explicitly exclude billing
    -- diagnoses, medical history, and problem-list diagnoses.
    WHERE def.Type = 'Encounter Diagnosis'

      -- Completed in-person outpatient encounter.
      -- The Caboodle face-to-face flag alone included telehealth/video visits,
      -- so also exclude video flags and virtual/phone VisitType labels.
      AND ef.DerivedEncounterStatus = 'Complete'
      AND ef.IsOutpatientFaceToFaceVisit = 1
      AND COALESCE(ef.IsVideoVisit, 0) = 0
      AND UPPER(COALESCE(ef.VisitType, '')) NOT LIKE '%TELEHEALTH%'
      AND UPPER(COALESCE(ef.VisitType, '')) NOT LIKE '%VIRTUAL%'
      AND UPPER(COALESCE(ef.VisitType, '')) NOT LIKE '%PHONE%'

      -- Inclusive calendar-date window that remains correct when ef.Date
      -- contains a time component.
      AND ef.Date >= @AomStartDate
      AND ef.Date < DATEADD(DAY, 1, @AomEndDate)

      -- Younger than 26 on the encounter date. This direct birthday comparison
      -- excludes future birth dates and excludes patients on/after age 26.
      -- Approved leap-day convention: SQL Server treats February 28 as the
      -- birthday in a non-leap year; no validated patients were affected.
      AND p.BirthDate IS NOT NULL
      AND p.BirthDate <= CAST(ef.Date AS DATE)
      AND CAST(ef.Date AS DATE) < DATEADD(YEAR, 26, p.BirthDate)

      -- Valid analytic patient only. These safeguards changed neither the
      -- validated index cohort nor the look-back population.
      AND p.IsValid = 1
      AND p.Test = 0
      AND p.DurableKey > 0
)

/* END SHARED DEFINITION: definitions/AOM-ENCOUNTER.sql */
```

```sql
/* BEGIN SHARED DEFINITION: definitions/AOM-TYMPANOSTOMY.sql (AOM-TUBE v0.3)
   CTEs: aom_index_first_dates, procedure_event_tube_dates,
         surgical_event_tube_dates, tympanostomy_dates,
         first_aom_tympanostomy */

, aom_index_first_dates AS (
    SELECT
        ae.PatientDurableKey,
        MIN(ae.EncounterDate) AS FirstAomDate
    FROM aom_encounters AS ae
    WHERE ae.EncounterDate >= @StudyStartDate
      AND ae.EncounterDate < DATEADD(DAY, 1, @StudyEndDate)
    GROUP BY ae.PatientDurableKey
),
procedure_event_tube_dates AS (
    SELECT DISTINCT
        pef.PatientDurableKey,
        CAST(pef.ProcedureStartInstant AS DATE) AS TubeDate
    FROM dbo.ProcedureEventFact AS pef
    WHERE pef._IsDeleted = 0
      AND COALESCE(pef.IsUnsuccessfulAttempt, 0) = 0
      AND pef.ProcedureStartInstant >= @StudyStartDate
      AND pef.ProcedureStartInstant < DATEADD(DAY, 1, @FollowupEndDate)
      AND pef.ProcedureCodeSet IN ('HcpcsCode', 'CPT(R)')
      AND pef.ProcedureCode IN ('69433', '69436', '0583T')
),
surgical_event_tube_dates AS (
    SELECT DISTINCT
        spef.PatientDurableKey,
        surgery_date.DateValue AS TubeDate
    FROM dbo.SurgicalProcedureEventFact AS spef
    INNER JOIN dbo.ProcedureDim AS procedure_code
        ON procedure_code.ProcedureKey = spef.ProcedureCodeKey
    INNER JOIN dbo.DateDim AS surgery_date
        ON surgery_date.DateKey = spef.SurgeryDateKey
    WHERE spef._IsDeleted = 0
      AND spef.Performed = 1
      AND surgery_date.DateValue >= @StudyStartDate
      AND surgery_date.DateValue < DATEADD(DAY, 1, @FollowupEndDate)
      AND (
            procedure_code.CptCode IN ('69433', '69436', '0583T')
         OR procedure_code.HcpcsCode IN ('69433', '69436', '0583T')
         OR procedure_code.Code IN ('69433', '69436', '0583T')
      )
),

-- UNION removes duplicate patient/date representations across both sources.
tympanostomy_dates AS (
    SELECT PatientDurableKey, TubeDate
    FROM procedure_event_tube_dates

    UNION

    SELECT PatientDurableKey, TubeDate
```

```sql
    FROM surgical_event_tube_dates
),
first_aom_tympanostomy AS (
    SELECT
        first_aom.PatientDurableKey,
        first_aom.FirstAomDate,
        MIN(tube.TubeDate) AS FirstTubeDate,
        DATEDIFF(DAY, first_aom.FirstAomDate, MIN(tube.TubeDate))
            AS DaysFirstAomToFirstTube
    FROM aom_index_first_dates AS first_aom
    INNER JOIN tympanostomy_dates AS tube
        ON tube.PatientDurableKey = first_aom.PatientDurableKey
       AND tube.TubeDate >= first_aom.FirstAomDate
    GROUP BY
        first_aom.PatientDurableKey,
        first_aom.FirstAomDate
)

/* END SHARED DEFINITION: definitions/AOM-TYMPANOSTOMY.sql */

/* BEGIN TASK-SPECIFIC LOGIC: Task 8 */

, distribution AS (
    SELECT DISTINCT
        CAST(
            PERCENTILE_CONT(0.25) WITHIN GROUP (
                ORDER BY tube.DaysFirstAomToFirstTube
            ) OVER ()
            AS DECIMAL(10, 2)
        ) AS Q1Days,
        CAST(
            PERCENTILE_CONT(0.50) WITHIN GROUP (
                ORDER BY tube.DaysFirstAomToFirstTube
            ) OVER ()
            AS DECIMAL(10, 2)
        ) AS MedianDays,
        CAST(
            PERCENTILE_CONT(0.75) WITHIN GROUP (
                ORDER BY tube.DaysFirstAomToFirstTube
            ) OVER ()
            AS DECIMAL(10, 2)
        ) AS Q3Days,
        COUNT(*) OVER () AS PatientsIncluded
    FROM first_aom_tympanostomy AS tube
)
SELECT
    distribution.MedianDays,
    distribution.Q1Days,
    distribution.Q3Days,
    CAST(
        distribution.Q3Days - distribution.Q1Days
        AS DECIMAL(10, 2)
    ) AS IqrWidthDays,
    distribution.PatientsIncluded
```

```
FROM distribution;

/* END TASK-SPECIFIC LOGIC: Task 8 */
```

### Task 9. Complications

```
/*
EXECUTION RESULT
Run date: 2026-07-31 (America/Los_Angeles)
Command: uv run python query.py -f .\queries\09_aom_patients_with_complication.sql

ComplicationType  PatientCount  AomCohortPatients
any_complication          1253              11091
conjunctivitis             978              11091
tm_perforation             302              11091
mastoiditis                  3              11091
*/

/*
Task 9: Complications

Shared definition copied below:
- definitions/AOM-ENCOUNTER.sql
- Complication ascertainment is task-specific and system-wide.
*/

DECLARE @StudyStartDate  DATE = '2023-01-01';
DECLARE @StudyEndDate    DATE = '2024-12-31';
DECLARE @FollowupEndDate DATE = '2025-12-31';
DECLARE @AomStartDate    DATE = @StudyStartDate;
DECLARE @AomEndDate      DATE = @StudyEndDate;

/* BEGIN SHARED DEFINITION: definitions/AOM-ENCOUNTER.sql (AOM-ENC v1.0)
   CTEs: smch_pediatric_departments, aom_diagnosis_keys, aom_encounters */

-- Exact protocol-supplied SMCH primary care pediatric departments.
-- All 30 IDs mapped and had utilization during validation. Keep the explicitly
-- supplied ZZZ-named historical IDs; do not infer an additional active-status
-- restriction from the department name.
WITH smch_pediatric_departments AS (
    SELECT v.DepartmentEpicId
    FROM (VALUES
        ('10801100'), ('10802100'), ('10803100'), ('10804100'), ('10806100'),
        ('10807100'), ('10808100'), ('10809100'), ('10810100'), ('10814100'),
        ('10815100'), ('10816100'), ('10829100'), ('10830100'), ('10837100'),
        ('10838100'), ('10839100'), ('10842100'), ('10843100'), ('10847100'),
        ('10850105'), ('10855101'), ('10860100'), ('10861100'), ('10863100'),
        ('10864100'), ('10865102'), ('10867100'), ('10868001'), ('10873100')
    ) AS v(DepartmentEpicId)
),

-- Exact AOM ICD-10-CM inclusion:
```

```sql
-- H66.001-H66.019, H66.40-H66.43, and H66.90-H66.93.
-- Resolving DiagnosisKey first handles multiple terminology rows per code.
aom_diagnosis_keys AS (
    SELECT DISTINCT
        dt.DiagnosisKey
    FROM dbo.DiagnosisTerminologyDim AS dt
    WHERE dt.Type = 'ICD-10-CM'
      AND (
            dt.Value LIKE 'H66.00[1-9]'
         OR dt.Value LIKE 'H66.01[0-9]'
         OR dt.Value IN (
                'H66.40', 'H66.41', 'H66.42', 'H66.43',
                'H66.90', 'H66.91', 'H66.92', 'H66.93'
         )
      )
),

-- One row per patient and qualifying AOM encounter. Retaining EncounterKey and
-- EncounterDate supports recurrence, antibiotic linkage, and follow-up tasks.
aom_encounters AS (
    SELECT DISTINCT
        def.PatientDurableKey,
        def.EncounterKey,
        CAST(ef.Date AS DATE) AS EncounterDate,
        dd.DepartmentEpicId
    FROM dbo.DiagnosisEventFact AS def
    INNER JOIN aom_diagnosis_keys AS aom
        ON aom.DiagnosisKey = def.DiagnosisKey

    -- Require the diagnosis to be attached to a real encounter.
    INNER JOIN dbo.EncounterFact AS ef
        ON ef.EncounterKey = def.EncounterKey

    -- Apply the supplied clinic list through the encounter's historical
    -- DepartmentKey and stable DepartmentEpicId.
    INNER JOIN dbo.DepartmentDim AS dd
        ON dd.DepartmentKey = ef.DepartmentKey
    INNER JOIN smch_pediatric_departments AS smch
        ON smch.DepartmentEpicId = dd.DepartmentEpicId

    -- PatientDim is history-tracked. Use exactly one current patient record.
    INNER JOIN dbo.PatientDim AS p
        ON p.DurableKey = def.PatientDurableKey
       AND p.IsCurrent = 1

    -- Diagnosis source: encounter diagnosis only. Explicitly exclude billing
    -- diagnoses, medical history, and problem-list diagnoses.
    WHERE def.Type = 'Encounter Diagnosis'

      -- Completed in-person outpatient encounter.
      -- The Caboodle face-to-face flag alone included telehealth/video visits,
      -- so also exclude video flags and virtual/phone VisitType labels.
      AND ef.DerivedEncounterStatus = 'Complete'
      AND ef.IsOutpatientFaceToFaceVisit = 1
```

```
      AND COALESCE(ef.IsVideoVisit, 0) = 0
      AND UPPER(COALESCE(ef.VisitType, '')) NOT LIKE '%TELEHEALTH%'
      AND UPPER(COALESCE(ef.VisitType, '')) NOT LIKE '%VIRTUAL%'
      AND UPPER(COALESCE(ef.VisitType, '')) NOT LIKE '%PHONE%'

      -- Inclusive calendar-date window that remains correct when ef.Date
      -- contains a time component.
      AND ef.Date >= @AomStartDate
      AND ef.Date < DATEADD(DAY, 1, @AomEndDate)

      -- Younger than 26 on the encounter date. This direct birthday comparison
      -- excludes future birth dates and excludes patients on/after age 26.
      -- Approved leap-day convention: SQL Server treats February 28 as the
      -- birthday in a non-leap year; no validated patients were affected.
      AND p.BirthDate IS NOT NULL
      AND p.BirthDate <= CAST(ef.Date AS DATE)
      AND CAST(ef.Date AS DATE) < DATEADD(YEAR, 26, p.BirthDate)

      -- Valid analytic patient only. These safeguards changed neither the
      -- validated index cohort nor the look-back population.
      AND p.IsValid = 1
      AND p.Test = 0
      AND p.DurableKey > 0
)

/* END SHARED DEFINITION: definitions/AOM-ENCOUNTER.sql */

/* BEGIN TASK-SPECIFIC LOGIC: Task 9 */

, aom_cohort_first_dates AS (
    SELECT
        ae.PatientDurableKey,
        MIN(ae.EncounterDate) AS FirstAomDate
    FROM aom_encounters AS ae
    GROUP BY ae.PatientDurableKey
),
complication_diagnosis_keys AS (
    SELECT DISTINCT
        terminology.DiagnosisKey,
        CASE
            WHEN terminology.Value LIKE 'H10.[0-3]%'
                THEN 'conjunctivitis'
            WHEN terminology.Value LIKE 'H72.[0-9]%'
                THEN 'tm_perforation'
            WHEN terminology.Value LIKE 'H70.0%'
                THEN 'mastoiditis'
        END AS ComplicationType
    FROM dbo.DiagnosisTerminologyDim AS terminology
    WHERE terminology.Type = 'ICD-10-CM'
      AND (
            terminology.Value LIKE 'H10.[0-3]%'
         OR terminology.Value LIKE 'H72.[0-9]%'
         OR terminology.Value LIKE 'H70.0%'
      )
```

```sql
),
complication_events AS (
    SELECT DISTINCT
        diagnosis.PatientDurableKey,
        diagnosis.EncounterKey,
        CAST(encounter.Date AS DATE) AS ComplicationDate,
        complication.ComplicationType
    FROM dbo.DiagnosisEventFact AS diagnosis
    INNER JOIN complication_diagnosis_keys AS complication
        ON complication.DiagnosisKey = diagnosis.DiagnosisKey
    INNER JOIN dbo.EncounterFact AS encounter
        ON encounter.EncounterKey = diagnosis.EncounterKey
    WHERE diagnosis.Type = 'Encounter Diagnosis'
      AND encounter.Date >= @StudyStartDate
      AND encounter.Date < DATEADD(DAY, 1, @FollowupEndDate)
),
matched_complications AS (
    -- Conjunctivitis counts only on Day 0 through Day +7 after any index AOM.
    SELECT DISTINCT
        aom.PatientDurableKey,
        complication.ComplicationType
    FROM aom_encounters AS aom
    INNER JOIN complication_events AS complication
        ON complication.PatientDurableKey = aom.PatientDurableKey
       AND complication.ComplicationType = 'conjunctivitis'
       AND complication.ComplicationDate >= aom.EncounterDate
       AND complication.ComplicationDate
            < DATEADD(DAY, 8, aom.EncounterDate)

    UNION

    -- Perforation and mastoiditis count from first index AOM through follow-up.
    SELECT DISTINCT
        cohort.PatientDurableKey,
        complication.ComplicationType
    FROM aom_cohort_first_dates AS cohort
    INNER JOIN complication_events AS complication
        ON complication.PatientDurableKey = cohort.PatientDurableKey
       AND complication.ComplicationType IN ('tm_perforation', 'mastoiditis')
       AND complication.ComplicationDate >= cohort.FirstAomDate
),
complication_types AS (
    SELECT v.ComplicationType, v.SortOrder
    FROM (VALUES
        ('any_complication', 0),
        ('conjunctivitis', 1),
        ('tm_perforation', 2),
        ('mastoiditis', 3)
    ) AS v(ComplicationType, SortOrder)
),
complication_counts AS (
    SELECT
        'any_complication' AS ComplicationType,
        COUNT(DISTINCT matched.PatientDurableKey) AS PatientCount
```

```
    FROM matched_complications AS matched

    UNION ALL

    SELECT
        type_list.ComplicationType,
        COUNT(DISTINCT matched.PatientDurableKey) AS PatientCount
    FROM complication_types AS type_list
    LEFT JOIN matched_complications AS matched
        ON matched.ComplicationType = type_list.ComplicationType
    WHERE type_list.ComplicationType <> 'any_complication'
    GROUP BY type_list.ComplicationType
),
cohort_size AS (
    SELECT COUNT(*) AS AomCohortPatients
    FROM aom_cohort_first_dates
)
SELECT
    type_list.ComplicationType,
    counts.PatientCount,
    cohort.AomCohortPatients
FROM complication_types AS type_list
INNER JOIN complication_counts AS counts
    ON counts.ComplicationType = type_list.ComplicationType
CROSS JOIN cohort_size AS cohort
ORDER BY type_list.SortOrder;

/* END TASK-SPECIFIC LOGIC: Task 9 */
```

### Task 10. Treatment Failure

```
/*
EXECUTION RESULT
Run date: 2026-07-31 (America/Los_Angeles)
Command: uv run python query.py -f .\queries\10_antibiotic_treated_patients_with_treatment_failure.sql

PatientsWithTreatmentFailure:       446
AntibioticTreatedCohortPatients:   10623
PercentWithTreatmentFailure:        4.20
*/

/*
Task 10: Treatment failure

Shared definitions copied below:
- definitions/AOM-ENCOUNTER.sql
- definitions/AOM-ANTIBIOTIC.sql
- AOM-ANTIBIOTIC supplies the first antibiotic and the AOM encounter to which
  that order was deterministically attributed.
*/

DECLARE @StudyStartDate DATE = '2023-01-01';
```

```sql
DECLARE @StudyEndDate   DATE = '2024-12-31';
DECLARE @AomStartDate   DATE = @StudyStartDate;
DECLARE @AomEndDate     DATE = @StudyEndDate;

/* BEGIN SHARED DEFINITION: definitions/AOM-ENCOUNTER.sql (AOM-ENC v1.0)
   CTEs: smch_pediatric_departments, aom_diagnosis_keys, aom_encounters */

-- Exact protocol-supplied SMCH primary care pediatric departments.
-- All 30 IDs mapped and had utilization during validation. Keep the explicitly
-- supplied ZZZ-named historical IDs; do not infer an additional active-status
-- restriction from the department name.
WITH smch_pediatric_departments AS (
    SELECT v.DepartmentEpicId
    FROM (VALUES
        ('10801100'), ('10802100'), ('10803100'), ('10804100'), ('10806100'),
        ('10807100'), ('10808100'), ('10809100'), ('10810100'), ('10814100'),
        ('10815100'), ('10816100'), ('10829100'), ('10830100'), ('10837100'),
        ('10838100'), ('10839100'), ('10842100'), ('10843100'), ('10847100'),
        ('10850105'), ('10855101'), ('10860100'), ('10861100'), ('10863100'),
        ('10864100'), ('10865102'), ('10867100'), ('10868001'), ('10873100')
    ) AS v(DepartmentEpicId)
),

-- Exact AOM ICD-10-CM inclusion:
-- H66.001-H66.019, H66.40-H66.43, and H66.90-H66.93.
-- Resolving DiagnosisKey first handles multiple terminology rows per code.
aom_diagnosis_keys AS (
    SELECT DISTINCT
        dt.DiagnosisKey
    FROM dbo.DiagnosisTerminologyDim AS dt
    WHERE dt.Type = 'ICD-10-CM'
      AND (
            dt.Value LIKE 'H66.00[1-9]'
         OR dt.Value LIKE 'H66.01[0-9]'
         OR dt.Value IN (
                'H66.40', 'H66.41', 'H66.42', 'H66.43',
                'H66.90', 'H66.91', 'H66.92', 'H66.93'
         )
      )
),

-- One row per patient and qualifying AOM encounter. Retaining EncounterKey and
-- EncounterDate supports recurrence, antibiotic linkage, and follow-up tasks.
aom_encounters AS (
    SELECT DISTINCT
        def.PatientDurableKey,
        def.EncounterKey,
        CAST(ef.Date AS DATE) AS EncounterDate,
        dd.DepartmentEpicId
    FROM dbo.DiagnosisEventFact AS def
    INNER JOIN aom_diagnosis_keys AS aom
        ON aom.DiagnosisKey = def.DiagnosisKey

    -- Require the diagnosis to be attached to a real encounter.
```

```
    INNER JOIN dbo.EncounterFact AS ef
        ON ef.EncounterKey = def.EncounterKey

    -- Apply the supplied clinic list through the encounter's historical
    -- DepartmentKey and stable DepartmentEpicId.
    INNER JOIN dbo.DepartmentDim AS dd
        ON dd.DepartmentKey = ef.DepartmentKey
    INNER JOIN smch_pediatric_departments AS smch
        ON smch.DepartmentEpicId = dd.DepartmentEpicId

    -- PatientDim is history-tracked. Use exactly one current patient record.
    INNER JOIN dbo.PatientDim AS p
        ON p.DurableKey = def.PatientDurableKey
       AND p.IsCurrent = 1

    -- Diagnosis source: encounter diagnosis only. Explicitly exclude billing
    -- diagnoses, medical history, and problem-list diagnoses.
    WHERE def.Type = 'Encounter Diagnosis'

      -- Completed in-person outpatient encounter.
      -- The Caboodle face-to-face flag alone included telehealth/video visits,
      -- so also exclude video flags and virtual/phone VisitType labels.
      AND ef.DerivedEncounterStatus = 'Complete'
      AND ef.IsOutpatientFaceToFaceVisit = 1
      AND COALESCE(ef.IsVideoVisit, 0) = 0
      AND UPPER(COALESCE(ef.VisitType, '')) NOT LIKE '%TELEHEALTH%'
      AND UPPER(COALESCE(ef.VisitType, '')) NOT LIKE '%VIRTUAL%'
      AND UPPER(COALESCE(ef.VisitType, '')) NOT LIKE '%PHONE%'

      -- Inclusive calendar-date window that remains correct when ef.Date
      -- contains a time component.
      AND ef.Date >= @AomStartDate
      AND ef.Date < DATEADD(DAY, 1, @AomEndDate)

      -- Younger than 26 on the encounter date. This direct birthday comparison
      -- excludes future birth dates and excludes patients on/after age 26.
      -- Approved leap-day convention: SQL Server treats February 28 as the
      -- birthday in a non-leap year; no validated patients were affected.
      AND p.BirthDate IS NOT NULL
      AND p.BirthDate <= CAST(ef.Date AS DATE)
      AND CAST(ef.Date AS DATE) < DATEADD(YEAR, 26, p.BirthDate)

      -- Valid analytic patient only. These safeguards changed neither the
      -- validated index cohort nor the look-back population.
      AND p.IsValid = 1
      AND p.Test = 0
      AND p.DurableKey > 0
)

/* END SHARED DEFINITION: definitions/AOM-ENCOUNTER.sql */

/* BEGIN SHARED DEFINITION: definitions/AOM-ANTIBIOTIC.sql (AOM-ABX v0.3)
   CTEs: systemic_antibiotic_order_base, systemic_antibiotic_orders,
         aom_antibiotic_order_candidates, aom_antibiotic_orders,
```

```sql
     first_aom_antibiotic_per_patient, antibiotic_treated_aom_cohort */

, systemic_antibiotic_order_base AS (
    SELECT
        mo.MedicationOrderKey,
        mo.PatientDurableKey,
        mo.EncounterKey AS OrderEncounterKey,
        mo.OrderedInstant,
        LOWER(md.SimpleGenericName) AS NormalizedIngredientName
    FROM dbo.MedicationOrderFact AS mo
    INNER JOIN dbo.MedicationDim AS md
        ON md.MedicationKey = mo.MedicationKey
    INNER JOIN dbo.DepartmentDim AS department
        ON department.DepartmentKey = mo.DepartmentKey
    INNER JOIN smch_pediatric_departments AS clinic
        ON clinic.DepartmentEpicId = department.DepartmentEpicId
    WHERE mo.Type = 'Medications'
      AND mo.IsPending = 0
      AND mo._IsDeleted = 0
      AND mo.OrderedInstant >= @StudyStartDate

      -- Keep return-treatment orders needed by Task 10. AOM linkage below
      -- remains restricted to Day 0 through Day 3.
      AND mo.OrderedInstant < DATEADD(DAY, 15, @StudyEndDate)
      AND UPPER(md.TherapeuticClass) = 'ANTIBIOTICS'
      AND UPPER(md.Route) IN ('PO', 'INJ', 'IV', 'IM', 'SUBQ', 'SC')
),
systemic_antibiotic_orders AS (
    SELECT
        base.MedicationOrderKey,
        base.PatientDurableKey,
        base.OrderEncounterKey,
        base.OrderedInstant,
        CASE
            WHEN base.NormalizedIngredientName LIKE '%amoxicillin%clav%'
                THEN 'amoxicillin-clavulanate'
            WHEN base.NormalizedIngredientName LIKE 'amoxicillin%'
                THEN 'amoxicillin'
            WHEN base.NormalizedIngredientName LIKE 'cefdinir%' THEN 'cefdinir'
            WHEN base.NormalizedIngredientName LIKE 'clindamycin%' THEN 'clindamycin'
            WHEN base.NormalizedIngredientName LIKE 'ceftriaxone%' THEN 'ceftriaxone'
            ELSE base.NormalizedIngredientName
        END AS AntibioticAgent
    FROM systemic_antibiotic_order_base AS base
),
aom_antibiotic_order_candidates AS (
    SELECT
        antibiotic.MedicationOrderKey,
        antibiotic.PatientDurableKey,
        aom.EncounterKey AS AomEncounterKey,
        aom.EncounterDate AS AomEncounterDate,
        antibiotic.OrderEncounterKey,
        antibiotic.OrderedInstant,
        antibiotic.AntibioticAgent,
```

```
        ROW_NUMBER() OVER (
            PARTITION BY
                antibiotic.PatientDurableKey,
                antibiotic.MedicationOrderKey
            ORDER BY aom.EncounterDate DESC, aom.EncounterKey DESC
        ) AS AomAttributionRank
    FROM systemic_antibiotic_orders AS antibiotic
    INNER JOIN aom_encounters AS aom
        ON aom.PatientDurableKey = antibiotic.PatientDurableKey
       AND antibiotic.OrderedInstant >= CAST(aom.EncounterDate AS DATETIME)
       AND antibiotic.OrderedInstant
            < DATEADD(DAY, 4, CAST(aom.EncounterDate AS DATETIME))
),
aom_antibiotic_orders AS (
    SELECT
        candidate.MedicationOrderKey,
        candidate.PatientDurableKey,
        candidate.AomEncounterKey,
        candidate.AomEncounterDate,
        candidate.OrderEncounterKey,
        candidate.OrderedInstant,
        candidate.AntibioticAgent,
        CASE
            WHEN candidate.AntibioticAgent IN (
                'amoxicillin',
                'amoxicillin-clavulanate',
                'cefdinir',
                'clindamycin',
                'ceftriaxone'
            ) THEN candidate.AntibioticAgent
            ELSE 'other'
        END AS NamedAntibioticCategory,
        CASE
            WHEN candidate.AntibioticAgent = 'amoxicillin'
                THEN 'narrow-spectrum'
            WHEN candidate.AntibioticAgent IN (
                'amoxicillin-clavulanate',
                'cefdinir',
                'clindamycin',
                'ceftriaxone'
            ) THEN 'broad-spectrum'
            ELSE 'other'
        END AS SpectrumCategory
    FROM aom_antibiotic_order_candidates AS candidate
    WHERE candidate.AomAttributionRank = 1
),
first_aom_antibiotic_per_patient AS (
    SELECT
        linked.*,
        ROW_NUMBER() OVER (
            PARTITION BY linked.PatientDurableKey
            ORDER BY linked.OrderedInstant, linked.MedicationOrderKey
        ) AS FirstAntibioticRank
    FROM aom_antibiotic_orders AS linked
```

```sql
),
antibiotic_treated_aom_cohort AS (
    SELECT
        first_order.PatientDurableKey,
        first_order.AomEncounterKey AS IndexAomEncounterKey,
        first_order.AomEncounterDate AS IndexAomEncounterDate,
        first_order.MedicationOrderKey AS FirstMedicationOrderKey,
        first_order.OrderEncounterKey AS FirstAntibioticOrderEncounterKey,
        first_order.OrderedInstant AS FirstAntibioticOrderedInstant,
        first_order.AntibioticAgent AS FirstAntibioticAgent,
        first_order.NamedAntibioticCategory AS FirstNamedAntibioticCategory,
        first_order.SpectrumCategory AS FirstSpectrumCategory
    FROM first_aom_antibiotic_per_patient AS first_order
    WHERE first_order.FirstAntibioticRank = 1
)

/* END SHARED DEFINITION: definitions/AOM-ANTIBIOTIC.sql */

/* BEGIN TASK-SPECIFIC LOGIC: Task 10 */

, return_encounters AS (
    SELECT DISTINCT
        cohort.PatientDurableKey,
        cohort.IndexAomEncounterKey,
        cohort.IndexAomEncounterDate,
        cohort.FirstAntibioticAgent,
        encounter.EncounterKey AS ReturnEncounterKey,
        CAST(encounter.Date AS DATE) AS ReturnEncounterDate
    FROM antibiotic_treated_aom_cohort AS cohort
    INNER JOIN dbo.EncounterFact AS encounter
        ON encounter.PatientDurableKey = cohort.PatientDurableKey
       AND encounter.Date
            >= DATEADD(DAY, 2, cohort.IndexAomEncounterDate)
       AND encounter.Date
            < DATEADD(DAY, 15, cohort.IndexAomEncounterDate)
    INNER JOIN dbo.DepartmentDim AS department
        ON department.DepartmentKey = encounter.DepartmentKey
    INNER JOIN smch_pediatric_departments AS clinic
        ON clinic.DepartmentEpicId = department.DepartmentEpicId
    WHERE encounter.DerivedEncounterStatus = 'Complete'
      AND encounter.IsOutpatientFaceToFaceVisit = 1
      AND COALESCE(encounter.IsVideoVisit, 0) = 0
      AND UPPER(COALESCE(encounter.VisitType, '')) NOT LIKE '%TELEHEALTH%'
      AND UPPER(COALESCE(encounter.VisitType, '')) NOT LIKE '%VIRTUAL%'
      AND UPPER(COALESCE(encounter.VisitType, '')) NOT LIKE '%PHONE%'
),
treatment_failure_patients AS (
    SELECT DISTINCT
        return_visit.PatientDurableKey
    FROM return_encounters AS return_visit
    INNER JOIN systemic_antibiotic_orders AS return_antibiotic
        ON return_antibiotic.PatientDurableKey = return_visit.PatientDurableKey
       AND return_antibiotic.OrderEncounterKey
            = return_visit.ReturnEncounterKey
```

```
       AND return_antibiotic.AntibioticAgent
            <> return_visit.FirstAntibioticAgent
)
SELECT
    COUNT(failure.PatientDurableKey) AS PatientsWithTreatmentFailure,
    COUNT(*) AS AntibioticTreatedCohortPatients,
    CAST(
        100.0 * COUNT(failure.PatientDurableKey)
        / NULLIF(COUNT(*), 0)
        AS DECIMAL(6, 2)
    ) AS PercentWithTreatmentFailure
FROM antibiotic_treated_aom_cohort AS cohort
LEFT JOIN treatment_failure_patients AS failure
    ON failure.PatientDurableKey = cohort.PatientDurableKey;

/* END TASK-SPECIFIC LOGIC: Task 10 */
```
