## Supplement D. Reference Standard Answers for "Can a General-Purpose Coding Agent Analyze a Production Hospital Data Warehouse?"

**Adjudicated; run 2026-07-31.** *These are the finalized analyst answers used as the comparator; the head-to-head agreement appears in Table 2, the error taxonomy in Table 3, and patient-level overlap in Table 4.*

| # | Task | Analyst Reference Answer |
|---|---|---|
| 1 | Birth Month | 593,073 |
| 2 | AOM Cohort | 11,091 |
| 3 | Recurrent AOM | 495 |
| 4 | Prescribed Antibiotic | 10,623 of 11,091 |
| 5 | Antibiotic Frequency (of 10,623 treated) | amoxicillin 8,595 (80.91%); amoxicillin-clavulanate 2,177 (20.49%); cefdinir 1,384 (13.03%); clindamycin 13 (0.12%); ceftriaxone 187 (1.76%); other 573 (5.39%) |
| 6 | Antibiotic Spectrum Classification (of 10,623) | narrow 8,225 (77.43%); broad 2,068 (19.47%); other 330 (3.11%) |
| 7 | Tubes (Recurrent) | 55 of 495 |
| 8 | Median Days to Tube | 208 (IQR 108 to 362); n = 237 |
| 9 | Complications (of 11,091) | any 1,253; conjunctivitis 978; TM perforation 302; mastoiditis 3 |
| 10 | Treatment Failure | 446 of 10,623 (4.20%) |

*Task 5 categories can exceed 100% because a patient may receive more than one antibiotic; Task 6 assigns each patient one first-antibiotic category. Counts are for the 2026-07-31 warehouse refresh.*
