## Supplement E. Interactive Session Summar for "Can a General-Purpose Coding Agent Analyze a Production Hospital Data Warehouse?"

### Supplement E. Interactive Session Summary

**Interactive session, by task.** One guided pass per task at medium effort. Feedback rounds count analyst turns; the analyst never stated a target value or wrote SQL. Full feedback is in Supplement F.

| Task | Feedback Rounds | Agent Final Answer | Reference | Result |
|---|---|---|---|---|
| 2 AOM Cohort | 0 (unaided) | 11,091 | 11,091 | Exact |
| 10 Treatment Failure | 2 | 446 of 10,623 (4.20%) | 446 of 10,623 (4.20%) | Exact |
| 8 Median Days to Tube | 5 | n = 237; median 209; IQR 110 to 362 | n = 237; median 208; IQR 108 to 362 | Near miss (set correct, median off by 1 day) |
| 7 Tubes (Recurrent) | 5 | 97 of 495 | 55 of 495 | Miss (denominator correct, tubes overcounted) |
