## Supplement F. Analyst Feedback and Restated Definitions for "Can a General-Purpose Coding Agent Analyze a Production Hospital Data Warehouse?"

The analyst feedback given during the interactive session is reproduced verbatim below, exactly as typed. None of it states a target value or contains SQL; the analyst pointed out where the query departed from the written definition and, where a plain hint did not suffice, named a field or table. The cohort task required no feedback.

**Task 2, AOM cohort. No feedback.** The agent's first query already excluded video visits and returned the reference cohort of 11,091.

**Task 7, tympanostomy tubes among recurrent patients. Five rounds:**

1. Verify your query is searching all the correct places for tubes. Verify the dates for 3 episode sequence.
2. Correct for finding additional sources of tube data, but can you explain your logic for the date restriction on the 3 episode sequence?
3. If a patient has 3 episodes in 2022, and an AOM encounter in 2023 or 2024, should they be included? yes or no
4. Assume that the definition of AOM requires at least 3 episodes, not exactly 3.
5. update the results based on that

**Task 8, time from first AOM to first tube. Five rounds:**

1. check the inclusion criteria for patients, visits, and tubes.
2. include the surgical event fact table.
3. determine if there are any other places or types of code you missed. coalesce nullable fields
4. why did you add BillingTransactionFact? you should not have used BillingTransactionFact. accept HcpcsCode as well
5. check whether you should coalesce isdeleted and isunsuccessfulattempt

**Task 10, treatment failure. Two rounds:**

1. verify route and visit inclusion criteria
2. Medications administered through tubes (e.g., G-Tubes and NG-Tubes) are considered enteral administrations

At the close of each session the agent was asked to restate, in plain language and without SQL, the definition it had used. For the two tasks it answered correctly, the restatement matched the reference definition, including the exclusion of video visits for the cohort and the enteral-route inclusion for treatment failure. For the two tube tasks the restatement encoded the ascertainment errors described in the Results, counting billing charges as evidence of a tube on the recurrent-tube task and, on that task, broadening recurrence to any three qualifying episodes across the look-back window. The four restated definitions are available on request.
