## Supplement G. Definition Reuse (Propagation and Consistency Batches) for "Can a General-Purpose Coding Agent Analyze a Production Hospital Data Warehouse?"

Both batches ran at medium reasoning effort and tested whether a definition corrected with a human could be reused rather than re-derived. The propagation batch prepended the corrected AOM-cohort definition (the agent's restated definition from the interactive cohort task) to the five tasks that depend on the cohort and re-ran each three times. The consistency batch prepended each interacted task's own corrected definition and re-ran it five times, to test whether a locked definition is reproduced.

**Table G1. Propagation batch (corrected cohort definition prepended; five cohort-dependent tasks, three runs each).**

| Task | Reference | Exact (/3) | Cohort Denominator Held at 11,091? | Note |
|---|---|---|---|---|
| 3 Recurrent AOM | 495 of 11,091 | 3/3 | Yes | - |
| 4 Prescribed Antibiotic | 10,623 of 11,091 | 3/3 | Yes | - |
| 5 Antibiotic Frequency | see Supplement D | 2/3 | Yes | one run differed on antibiotic route (10,619) |
| 6 Antibiotic Spectrum Classification | see Supplement D | 3/3 | Yes | - |
| 9 Complications | see Supplement D | 2/3 | Yes | one run differed on conjunctivitis ascertainment |
| **Total** | | **13/15** | **15/15** | residual misses are task-specific logic the cohort definition did not address |

**Table G2. Consistency batch (each task's own corrected definition prepended; four interacted tasks, five runs each).**

| Task | Reference | Answers Across Five Runs | Exact (/5) | Reproducible Across Runs? |
|---|---|---|---|---|
| 2 AOM Cohort | 11,091 | 11,091 in all five | 5/5 | Yes (reproducible and correct) |
| 10 Treatment Failure | 446 of 10,623 | 446 of 10,623 in all five | 5/5 | Yes (reproducible and correct) |
| 8 Median Days to Tube | median 208, n = 237 | n = 237 in all five; median 208 in four, 209 in one | 4/5 | Set reproducible; median varies by one day |
| 7 Tubes (Recurrent) | 55 of 495 | 97 of 495 in all five | 0/5 | Reproducible but wrong (reproduces the billing-charge overcount) |

*Propagation shows that fixing one shared definition corrected the dependent tasks (13 of 15 exact) and eliminated the cohort drift (denominator held in all 15 runs). Consistency shows that locking a definition made the answer reproducible on every task, but correct only where the supplied definition was itself correct.*
