## Supplement H. SQUIRE 2.0 Checklist for "Can a General-Purpose Coding Agent Analyze a Production Hospital Data Warehouse?"

We report this study against three complementary guidelines: SQUIRE 2.0 (quality-improvement framing), TRIPOD-LLM (evaluation of a large language model), and STARD 2015 (accuracy against a reference standard). For STARD, the index test is the autonomous coding agent and the reference standard is the two-analyst adjudicated answer. TRIPOD-LLM items specific to de novo model development, fine-tuning, annotation, summarization, or translation are marked not applicable because we evaluated an existing general-purpose model. Section references point to the relevant part of this manuscript.

| **Section** | **Item** | **Checklist Item (Abbreviated)** | **Reported In** |
|---|---|---|---|
| Title | 1 | Indicates the report concerns improvement in healthcare | Title |
| Abstract | 2 | Summarizes the aim, methods, results, and conclusions | Abstract |
| Introduction | 3 | Problem description: nature and significance of the local problem | Introduction |
| Introduction | 4 | Available knowledge: what is already known, with citations | Introduction |
| Introduction | 5 | Rationale: concepts/frameworks used to explain the problem and intervention | Introduction; Discussion |
| Introduction | 6 | Specific aims of the project | Introduction (objective) |
| Methods | 7 | Context: contextual elements relevant to the work | Methods (Design, setting, governance; Data environment) |
| Methods | 8 | Intervention(s): description sufficient for others to reproduce | Methods (Access approach; Index method; Tasks and conditions); Supplement A, B; Figure 1 |
| Methods | 9 | Study of the intervention(s): approach to assessing impact | Methods (four conditions; Reference standard; Outcomes and analysis) |
| Methods | 10 | Measures: measures chosen, operational definitions, and their validity | Methods (Outcomes and analysis); Tables 1–4 |
| Methods | 11 | Analysis: qualitative and quantitative methods | Methods (Outcomes and analysis) |
| Methods | 12 | Ethical considerations | Methods (QI/NHSR determination #88354; AI Oversight Committee; BAA and privacy review) |
| Results | 13 | Results: intervention delivery, process/outcome data, unintended consequences, missing data | Results (implementation comparison; autonomous, assumption-listing, interactive, and reuse conditions: validity, accuracy, reproducibility, error taxonomy, cost, patient-level F1) |
| Discussion | 14 | Summary: key findings and strengths | Discussion (principal finding) |

| Section | Item | Checklist Item (Abbreviated) | Reported In |
|---|---|---|---|
| Discussion | 15 | Interpretation: association with intervention, comparison with literature, tradeoffs | Discussion |
| Discussion | 16 | Limitations | Discussion (limitations) |
| Discussion | 17 | Conclusions: usefulness, implications, next steps | Discussion (conclusion) |
| Other | 18 | Funding | Statements (Funding) |
