## Supplement I. TRIPOD-LLM Checklist for "Can a General-Purpose Coding Agent Analyze a Production Hospital Data Warehouse?"

| Section | Item | Checklist Item (Abbreviated) | Reported In |
|---|---|---|---|
| Title | 1 | Identify the study as evaluating an LLM, with task, population, and outcome | Title |
| Abstract | 2 | Structured abstract (see TRIPOD-LLM for Abstracts) | Abstract |
| Introduction | 3a | Healthcare context/use case and rationale, with references | Introduction |
| Introduction | 3b | Target population, intended use in the care pathway, and intended users | Introduction; Methods (setting; users = analysts) |
| Introduction | 4 | Objectives, including whether development, fine-tuning, or validation | Introduction (evaluation of an existing model) |
| Methods (Data) | 5a | Data sources for training/tuning/evaluation and rationale | Methods (Data environment); model training data proprietary/not available |
| Methods (Data) | 5b | Data points and description of their distribution | Methods (Data environment); Table 1 |
| Methods (Data) | 5c | Dates of oldest and newest text used | Methods (index window 2023–2024; snapshot = run-day refresh); model training dates not available |
| Methods (Data) | 5d | Data pre-processing and quality checking | Methods (Data environment; reporting-optimized warehouse) |
| Methods (Data) | 5e | Handling of missing and imbalanced data | Methods (Outcomes and analysis) |
| Methods (Analytical) | 6a | LLM name, version, and last training date | Methods (Index method: OpenAI Codex, GPT-5.6-sol); last training date not available (closed model) |
| Methods (Analytical) | 6b | LLM architecture, training, fine-tuning, alignment | N/A (existing closed model; not available) |
| Methods (Analytical) | 6c | Text-generation details, prompt engineering, inference settings | Methods (Index method: minimal system prompt; extra-high and medium reasoning; tools disabled); Supplement A; seed/temperature not exposed |
| Methods (Analytical) | 6d | Initial and post-processed output of the LLM | Methods (agent reports its answer and SQL) |
| Methods (Analytical) | 6e | Classification/probabilities/thresholds | N/A for classification; tolerance thresholds (±5%) in Methods (Outcomes and analysis) |
| LLM Output | 7a | Metrics capturing output quality vs reference standards | Methods (Outcomes: accuracy vs reference; SQL validity) |

| Section | Item | Checklist Item (Abbreviated) | Reported In |
|---|---|---|---|
| LLM Output | 7b | Relevance of metrics to the downstream task; correlation to human evaluation | Methods; Discussion (human-adjudicated standard) |
| LLM Output | 7c | Define the outcome, how predictions are calculated, date of inference, metrics | Methods (Outcomes and analysis); inference on each run's nightly snapshot; reference run 2026-07-31 |
| LLM Output | 7d | For subjective assessment: assessor qualifications, instructions, agreement | Methods (Reference standard: two analysts, adjudication; two pediatric ID physicians) |
| LLM Output | 7e | How performance was compared to other LLMs, humans, benchmarks | Methods (head-to-head vs analysts; medium vs extra-high reasoning) |
| Annotation | 8a–8c | Annotation labeling, annotators, and background | N/A (no dataset annotation) |
| Prompting | 9a | Prompt design, curation, and selection | Methods (calibration; prompt-lock); Supplement B |
| Prompting | 9b | Data used to develop the prompts | Methods (calibration set) |
| Summarization | 10 | Preprocessing before summarization | N/A (not a summarization task) |
| Instruction tuning | 11 | Instruction-tuning/alignment strategy and evaluation | N/A (no tuning) |
| Compute | 12 | Compute or proxies (time, cost, inference time, FLOPs) | Methods (Outcomes: time and tokens); Results (Cost: median duration, tokens, query iterations by setting) |
| Ethical approval | 13 | Ethics committee approval and consent or waiver | Methods (QI/NHSR determination #88354) |
| Open science | 14a | Funding source and role of funders | Statements (Funding) |
| Open science | 14b | Conflicts of interest and financial disclosures | Statements (Conflicts of interest) |
| Open science | 14c | Where the study protocol can be accessed | Protocol operationalized in Supplements A, B, and C |
| Open science | 14d | Study registration or statement of none | Statements (Registration): not registered; quality-improvement activity |
| Open science | 14e | Availability of study data | Statements (Data availability) |
| Open science | 14f | Availability of code to reproduce results | Statements (Code availability); Supplements A, B, and C |
| Public involvement | 15 | Patient/public involvement or statement of none | Statements (Patient and public involvement): none |
| Results (Participants) | 16a | Flow of data through the study | Results (per-task counts); Supplement D. No participant-flow diagram; the analytic unit is the task, not enrolled patients |

| Section | Item | Checklist Item (Abbreviated) | Reported In |
|---|---|---|---|
| Results (Participants) | 16b | Characteristics by source/setting; key dates; sample size | Methods (Data environment; index window 2023–2024); Results (cohort denominators). No patient-level baseline; the study compares query results, not patients |
| Results (Participants) | 16c | Distribution of clinical variables, development vs evaluation | N/A (no development/evaluation split; single warehouse) |
| Results (Participants) | 16d | Participants and outcome events per analysis | Results (cohort denominators; patient-level agreement, Table 4) |
| Performance | 17 | Performance per pre-specified metrics and/or human evaluation | Results (exact agreement, tolerance accuracy, reproducibility, error taxonomy, interactive session, and patient-level precision, recall, and F1; Tables 2 and 4, and Supplement E) |
| LLM updating | 18 | Results of any LLM updating | N/A (model pinned; no updating) |
| Discussion | 19a | Overall interpretation, including fairness | Discussion |
| Discussion | 19b | Limitations, biases, uncertainty, generalizability | Discussion (limitations) |
| Discussion | 19c | Challenges in using the data (representation, missingness, bias) | Discussion; Methods |
| Discussion | 19d | Intended use: input, end-user, autonomy/human oversight | Discussion (analyst-supervised use) |
| Discussion | 19e | Handling poor/unavailable input; usability in clinical care | Discussion (not clinician-facing; analyst validation) |
| Discussion | 19f | User interaction and expertise required | Discussion (interactive condition; oversight) |
| Discussion | 19g | Next steps for future research | Discussion (conclusion) |
