## Supplement J. STARD 2015 Checklist for "Can a General-Purpose Coding Agent Analyze a Production Hospital Data Warehouse?"

*Index test = the autonomous coding agent; reference standard = the two-analyst adjudicated answer. This is an analytic-accuracy comparison, not a clinical diagnostic-accuracy study, so several diagnostic-specific items are not applicable and are marked as such.*

| Section | Item | Checklist Item (Abbreviated) | Reported In |
|---|---|---|---|
| Title/Abstract | 1 | Identification as an accuracy study with a measure of accuracy | Title; Abstract (agent vs reference; F1, tolerance) |
| Abstract | 2 | Structured summary of design, methods, results, conclusions | Abstract |
| Introduction | 3 | Background, including intended use and role of the index test | Introduction |
| Introduction | 4 | Study objectives and hypotheses | Introduction |
| Methods | 5 | Prospective vs retrospective data collection | Methods (retrospective; existing operational data) |
| Methods | 6 | Eligibility criteria | Methods (Tasks and conditions); Supplement B; Table 1 |
| Methods | 7 | Basis on which eligible cases were identified | Supplement B (diagnosis codes, encounter type) |
| Methods | 8 | Where and when cases were identified (setting, dates) | Methods (SMCH outpatient; 2023–2024 index window) |
| Methods | 9 | Consecutive, random, or convenience series | Methods (all eligible encounters, i.e., complete series) |
| Methods | 10a | Index test in sufficient detail to replicate | Methods (Index method); Supplement A; Figure 1 |
| Methods | 10b | Reference standard in sufficient detail to replicate | Methods (Reference standard) |
| Methods | 11 | Rationale for the reference standard | Methods (Reference standard; automated metrics inadequate) |
| Methods | 12a | Index-test result categories/cut-offs (pre-specified vs exploratory) | Methods (exact match; ±5% tolerance) |
| Methods | 12b | Reference-standard categories/cut-offs | Methods (adjudicated answer) |
| Methods | 13a | Whether reference results were available to index-test performers | Methods (agent run independently, before unblinding) |
| Methods | 13b | Whether index results were available to reference assessors | Methods (analysts blinded; adjudication before unblinding) |
| Methods | 14 | Methods for estimating/comparing accuracy | Methods (Outcomes: precision/recall/F1; clustered by question) |
| Methods | 15 | Handling of indeterminate results | Methods (SQL validity): no indeterminate results; all 60 runs executed and returned a value (Results) |
| Methods | 16 | Handling of missing data | Methods (Outcomes and analysis) |

| Section | Item | Checklist Item (Abbreviated) | Reported In |
|---|---|---|---|
| Methods | 17 | Analyses of variability (pre-specified vs exploratory) | Methods (triplicate reproducibility; medium vs extra-high) |
| Methods | 18 | Intended sample size and how determined | Methods (feasibility; ~10 tasks; descriptive, no formal power) |
| Results | 19 | Flow of cases (diagram) | Figure 1 (study design); Results (per-task cohort counts). Not a patient-flow study; no case-flow diagram applies |
| Results | 20 | Baseline characteristics | N/A: accuracy comparison of query results, not a patient cohort with baseline characteristics |
| Results | 21a/21b | Severity/alternative-diagnosis distribution | N/A (no disease-classification target) |
| Results | 22 | Time interval/interventions between index test and reference | N/A (both computed from the same frozen data snapshot) |
| Results | 23 | Cross-tabulation of index vs reference results | Results/Tables 2–3 (per-task agreement; not a 2×2, not binary) |
| Results | 24 | Estimates of accuracy and precision (95% CI) | Results (precision, recall, F1; Table 4); 95% CIs not reported (descriptive feasibility analysis) |
| Results | 25 | Adverse events from index test or reference standard | N/A (read-only analytics; no patient testing) |
| Discussion | 26 | Limitations, bias, uncertainty, generalizability | Discussion (limitations) |
| Discussion | 27 | Implications for practice; intended use/role | Discussion (analyst-supervised use) |
| Other | 28 | Registration number and registry | Statements (Registration): not registered; quality-improvement activity |
| Other | 29 | Where the full protocol can be accessed | Supplements A, B, and C |
| Other | 30 | Funding sources and role of funders | Statements (Funding) |
